# Neural, Behavioral, and Clinical Outcomes of a Voice-based AI Coach for Depression and Anxiety: A Phase 2 Randomized Trial

**DOI:** 10.64898/2025.12.22.25342792

**Authors:** Thomas Kannampallil, Olusola A. Ajilore, Joshua M. Smyth, Amruta Barve, Corina R. Ronneberg, Nan Lv, Vikas Kumar, Claudia Garcia, Gbenga Aborisade, Nancy E. Wittels, Zhengxin Tang, Bharathi Chinnakotla, Lan Xiao, Jun Ma

**Author notes:** Corresponding Author: Jun Ma, MD, PhD, Beth and George Vitoux Professor of Medicine, Founding director, Vitoux Program on Aging and Prevention, Department of Medicine, University of Illinois at Chicago, 1747 W. Roosevelt Rd, Room 586 (MC 275), Chicago, IL 60608. Equal first authors (Thomas Kannampallil, Olusola A. Ajilore).

## Abstract

**Importance:** Artificial Intelligence (AI) voice-based chatbots may improve access to evidence-based psychotherapy for depression and anxiety, but their therapeutic mechanisms and outcomes are largely unknown.

**Objective:** To investigate the mechanisms and potential efficacy of Lumen, an AI voice-based coach delivering problem-solving treatment (PST), among adults with untreated, clinically significant depression and/or anxiety.

**Design:** Phase 2, 3-arm randomized clinical trial.

**Setting:** A public university and affiliated medical center in Chicago, Illinois.

**Participants:** Adults with a Patient Health Questionnaire-9 score of 10-19 and/or a Generalized Anxiety Disorder Scale score of 10-14 were randomized to Lumen-coached PST (n=100), human-coached PST (n=50), or waitlist control (n=50).

**Interventions:** PST was delivered in 4 weekly sessions followed by 4 biweekly sessions, by Lumen—a custom-developed, rule-based AI application on Amazon’s Alexa platform—or by a human coach via videoconferencing.

**Main Outcome(s) and Measure(s):** The primary mechanistic outcome was change in right dorsolateral prefrontal cortex (dlPFC) activation for cognitive control, assessed with functional neuroimaging. Secondary outcomes were patient-reported, theory-based behavioral targets (e.g., problem-solving) and clinical symptoms (e.g., psychological distress).

**Results:** Among 200 participants (mean (SD) age, 36.6 (11.9) years; 77% women; 25% Black American; 28% Latino), change from baseline in right dlPFC activity at 18 weeks did not differ significantly by treatment arm. Compared with waitlist control, Lumen-coached participants had significantly greater improvements from baseline to 18 weeks in overall problem-solving ability (mean difference, 1.04; 95% CI, 0.23—1.84]) and psychological distress (mean difference, −3.56; 95% CI −5.69 to −1.43) due to depressive and anxiety symptoms. Lumen- and human-coached PST did not differ significantly on either measure. One serious adverse event (hospitalization), unrelated to this study, was reported.

**Conclusions and Relevance:** In this phase 2 experimental-therapeutics RCT, neither AI- nor human-coached PST significantly engaged the prespecified neural target for cognitive control. However, both interventions improved patient-reported problem-solving and psychological distress relative to waitlist control, providing exploratory evidence of a non-neural candidate mechanism and a clinical signal warranting future testing.

**Trial Registration:** clinicaltrials.gov, <u>NCT05603923</u>

**Key Points:** *Question:* Does Lumen, an Artificial Intelligence (AI) voice-based coach delivering problem-solving treatment (PST) for depression and anxiety, engage a mechanistic neural target for cognitive control (right dorsolateral prefrontal cortex, dlPFC) better than waitlist, and not worse than human-coached PST, and, secondarily, does it improve problem-solving behavior and clinical symptoms?

*Findings:* In this phase 2, 3-arm experimental-therapeutics randomized trial among 200 adults with untreated, clinically significant depression and/or anxiety, neither Lumen- nor human-coached PST significantly engaged the neural target versus waitlist control. Both interventions, however, significantly improved secondary outcomes related to patient-reported problem-solving ability and psychological distress due to depressive and anxiety symptoms relative to waitlist control.

*Meaning:* Despite null neural-target findings, Lumen may hold promise as a scalable intervention for improving problem-solving and psychological distress in adults with depression and/or anxiety.

## Introduction

Depression and anxiety are among the most common mental health disorders in the United States^1^ and leading causes of disability worldwide,^2^ and the two conditions frequently coexist. Yet fewer than half of individuals with symptoms receive treatment,^3,4^ and access to evidence-based psychotherapies remains limited by clinician shortages, stigma, and cost.^5,6^

Digital mental health tools, ranging from mobile applications to artificial intelligence (AI), offer alternative modalities for delivering psychotherapy. The most common digital AI interventions are chatbots, which use content delivery frameworks ranging from rule-based systems to large language models;^7^ however, evidence for their effects on depressive and anxiety symptoms remains mixed.^7–10^ Compared with text-based chatbots, voice-based interactions enable more natural, human-like conversation,^11,12^ though clinical research on this modality remains nascent.^13–15^

Developed through iterative design and formative usability testing,^16,17^ Lumen is an AI voice-based coach that uses rule-based natural language processing (NLP) to deliver problem-solving treatment (PST),^18^ a clinically-proven, cognition-targeted psychotherapy for depression and anxiety. In a phase 1 trial among adults with clinically significant depression and/or anxiety, Lumen-coached PST meaningfully engaged the a priori neural mechanistic target for cognitive control—right dorsolateral prefrontal cortex (dlPFC)—and produced symptom improvements meeting the prespecified criteria for additional testing.^19^

Guided by the experimental therapeutics framework, which prioritizes mechanism-targeted intervention testing,^20^ this larger phase 2, 3-arm randomized trial^21^ was designed to confirm that Lumen-coached PST was superior to waitlist control in engaging the right dlPFC for cognitive control—the primary mechanistic target. Secondary aims were to examine (1) whether Lumen-coached PST was noninferior to human-coached PST on right dlPFC engagement; (2) the effects of Lumen-coached PST, relative to waitlist control and human-coached PST, on additional prespecified neural and patient-reported measures of target engagement and on patient-reported symptom and functional outcomes; and (3) the mediating effects of target engagement on outcomes.

## Methods

### Participants

The trial was registered (Clinicaltrials.gov NCT05603923), and the protocol was published.^21^ Participants were enrolled between January 23, 2023, and December 17, 2024, through outpatient clinics at the University of Illinois Hospital and Health Sciences System and email listservs at the University of Illinois Chicago (Supplement, Section A).

Adults aged ≥18 reporting moderate or moderately-severe depressive symptoms (Patient Health Questionnaire [PHQ-9] scores 10–19)^22^ and/or moderate anxiety symptoms (Generalized Anxiety Disorder [GAD-7] scores 10–14)^23^ were eligible. Main exclusion criteria included serious medical or psychiatric comorbidities, concurrent psychiatric treatment, and suicidality (see full list in Supplement, Section B).

### Randomization and masking

Participants were randomized to Lumen-coached PST, human-coached PST, or waitlist control using a validated online system^24^ based on covariate-adaptive, biased-coin minimization.^25^ This approach achieved better-than-chance marginal balance across study arms on key baseline characteristics: sex, age, race/ethnicity, digital health literacy,^26^ PHQ-9 score, and GAD-7 score. Investigators, outcome assessors, and the analyst and safety monitor were masked to treatment assignment.

### Interventions

#### Lumen-coached PST

Developed on Amazon’s Alexa platform, Lumen delivered 8 PST sessions (4 weekly, then 4 biweekly). PST is patient-driven; its inherent structure and pragmatism are well-suited for AI voice-based delivery. Lumen guides participants to identify a problem, set a goal, brainstorm and compare solutions, choose a solution, develop an action plan, and implement and evaluate the plan in each session.^18^

Lumen uses rule-driven, retrieval-based NLP techniques to align with PST’s stepwise, algorithmic structure, guarding against hallucinations and off-topic conversation. Lumen previously demonstrated feasibility, treatment fidelity, and usability,^27^ along with promising neural target engagement and clinical benefit in a phase 1 trial^19^ (Supplement, Sections C-E).

Participants accessed Lumen using the Alexa app on a study-provided iPad and initiated each session with a voice command (“Alexa, Open Lumen, Session [number]).” Between sessions, participants completed surveys and ecological momentary assessments (EMAs) and received session reminders.

#### Human-coached PST

A doctoral-level, PST-certified health coach, who had previously delivered in-person PST in another trial,^28^ conducted the first session in person and the remaining 3 weekly and 4 biweekly sessions via Zoom. Session structure and content followed the PST manual^18^ and paralleled Lumen-coached sessions. Monthly fidelity evaluations by a study-independent master PST trainer identified no need for corrective actions.

#### Waitlist control

Waitlist control participants completed surveys and EMAs at intervals similar to those of the Lumen- and human-coached arms during the intervention period and could opt to receive a Lumen-enabled iPad after completing end-of-study assessments.

### Measures

Blinded outcome assessors conducted standardized assessments at baseline and 18 weeks, including neural and patient-reported mechanistic outcomes related to cognitive control and emotion reactivity, as well as clinical and functional outcomes.

#### Primary outcome

Task-based functional magnetic resonance imaging (fMRI) data were acquired using established sequences and parameters (Supplement, Section F). Informed by phase 1 findings,^19,28,29^ the primary region of interest (ROI) was the right dlPFC, engaged via a go/no-go task for cognitive control.

#### Secondary outcomes

##### Neural mechanistic targets

Secondary ROIs included the left dlPFC for cognitive control and bilateral amygdala activation during non-conscious viewing of threat and sad faces (negative affect), as a measure of emotion reactivity. Person-level activation for each task contrast of interest (e.g., no-go vs go, threat vs neutral faces) was derived consistent with prior studies.^30,31^

##### Patient-reported mechanistic targets

As a measure of cognitive control-associated behavior and a plausible, theory-based mediator of PST’s effects on symptom outcomes, the Social Problem-solving Inventory-Revised Short Form (SPSI-R:S) assessed overall problem-solving ability and 5 subscales: problem orientation (positive [PPO] and negative [NPO]) and problem-solving styles (rational [RPS], impulsive/careless [ICS], and avoidant [AS]).^32^ Additional patient-reported measures of cognitive control and emotion reactivity included the Dysfunctional Attitudes Scale (DAS),^33^ Penn State Worry Questionnaire (PSWQ),^34^ and Positive and Negative Affect Schedule (PANAS).^35^

##### Symptom and functional outcomes

The Hospital Anxiety and Depression Scale (HADS)^36,37^ measures depressive and anxiety symptom scores and a summative psychological distress score. The Sheehan Disability Scale (SDS)^38^ and the Work Productivity and Activity Impairment (WPAI) questionnaire^39^ assessed functional impairment (see also the protocol^21^).

### Statistical analysis

We conducted intention-to-treat (ITT) analyses for superiority tests (Lumen-coached and human-coached vs waitlist control) and both ITT and per-protocol analyses (i.e., excluding participants who did not complete all 8 sessions) for noninferiority tests (Lumen-coached vs human-coached).^40^ Analysis of between-group differences included all participants with 18-week data for each outcome, based on randomized group assignment.

Because neural target activation was expressed in standard deviation (SD) units relative to previously validated healthy control norms,^41,42^ between-group differences in change from baseline to 18 weeks were assessed using Student *t* tests.

Between-group differences in change in patient-reported outcomes from baseline to 18 weeks were tested using analysis of covariance (ANCOVA), assuming normality of error terms and adjusting for baseline values of the outcome measure. We reported model-adjusted between-group mean differences with 95% confidence intervals (CIs) and 2-tailed P values for superiority tests, and with 90% CIs and 1-tailed P values for noninferiority tests. To correct for multiple comparisons, we additionally reported P values adjusted for the false discovery rate (FDR). Statistical significance was set at α=.05. Cohen d was calculated using the mean difference between two groups divided by the pooled SD. A Cohen d of 0.3 (small effect) was the prespecified noninferiority margin for comparing Lumen- and human-coach PST on change in right dlPFC activation. Noninferiority margins were not prespecified for secondary outcomes given their exploratory nature and the focus on effect estimation to inform future research.

Using the approach of Kraemer et al,^43^ we defined mediation as present if a potential mediator met two conditions: first, the effect of Lumen- or human-coached PST vs waitlist control (X) on the potential mediator (M; path A, X→M) was statistically significant; second, the potential mediator was significantly associated with the outcome (Y), either as a main effect or as an interaction effect with either treatment vs waitlist control (path B, M→Y). Because all data were assessed at baseline and 18 weeks only, the focus was on contemporaneous mediation.^44^

All analyses were conducted using SAS, version 9.4 (SAS Institute Inc).

### Sample size calculation

To obtain additional data on Lumen as the experimental treatment and inform its further development, we used a 2:1:1 allocation to the Lumen-coached, human-coached, and waitlist control arms. The primary analysis was a superiority test of change in the primary neural target (right dlPFC) comparing Lumen-coached PST with waitlist control. A sample of 150 (100 Lumen, 50 waitlist control) was sufficient to provide 80% power to detect a medium effect (Cohen d = 0.45) at α=.05 (2-sided), assuming 85% retention. The human-coached and waitlist control arms were designed to be of equal size, for a total sample of 200.

## Results

### Sample characteristics and retention

Of 3176 individuals screened, 2773 were ineligible and 142 declined or were unable to participate (Figure 1).

**Figure 1.**
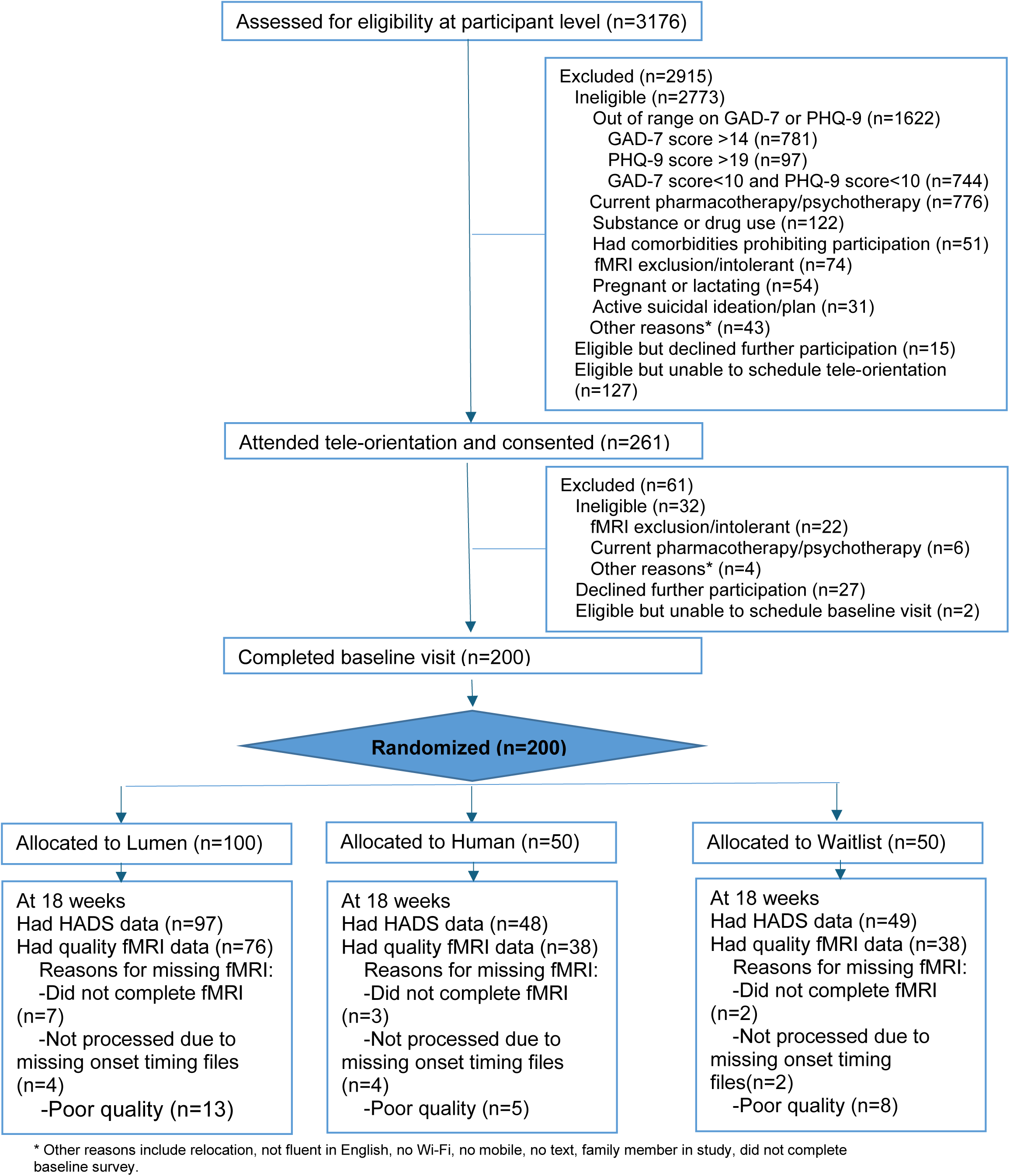
Consort chart.

Participants (N=200) had a mean (SD) age of 36.6 (11.9) years and included 77.5% women, 25.0% Black American, and 28.5% Latino; 62.0% had not completed postcollege education, and 47.5% had an annual family income below $55,000 (Table 1). At baseline, mean (SD) PHQ-9 and GAD-7 scores were 12.3 (3.5) and 10.9 (2.4), respectively, indicating moderate symptom severity on average; 61.5% of participants had PHQ-9 and GAD-7 scores ≥10, indicating clinically significant symptoms of both depression and anxiety.

**Table 1.**
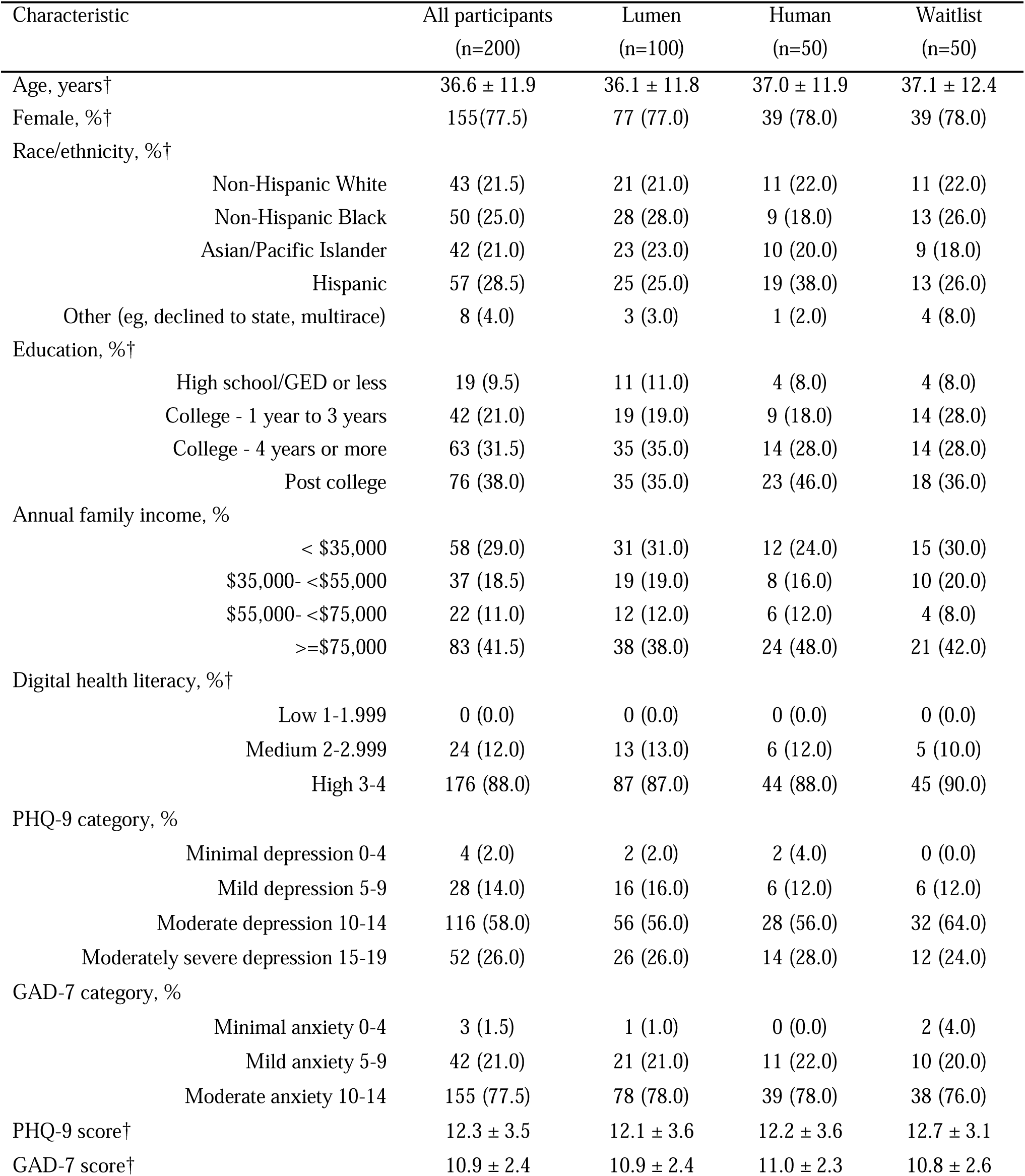
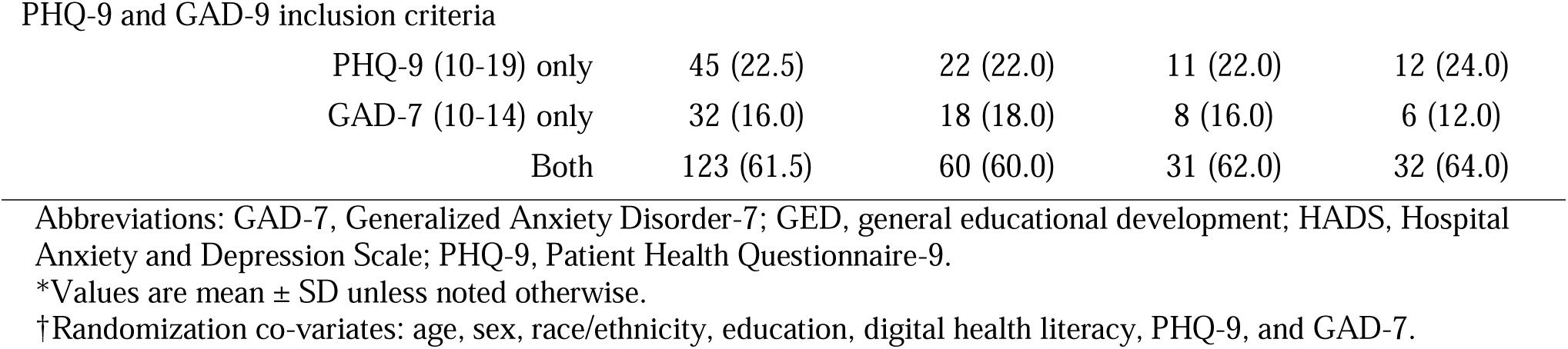
Baseline characteristics*.

| Characteristic | All participants<br>(n=200) | Lumen<br>(n=100) | Human<br>(n=50) | Waitlist<br>(n=50) |
| --- | --- | --- | --- | --- |
| Age, years† | 36.6 ± 11.9 | 36.1 ± 11.8 | 37.0 ± 11.9 | 37.1 ± 12.4 |
| Female, %† | 155(77.5) | 77 (77.0) | 39 (78.0) | 39 (78.0) |
| Race/ethnicity, %† |  |  |  |  |
| Non-Hispanic White | 43 (21.5) | 21 (21.0) | 11 (22.0) | 11 (22.0) |
| Non-Hispanic Black | 50 (25.0) | 28 (28.0) | 9 (18.0) | 13 (26.0) |
| Asian/Pacific Islander | 42 (21.0) | 23 (23.0) | 10 (20.0) | 9 (18.0) |
| Hispanic | 57 (28.5) | 25 (25.0) | 19 (38.0) | 13 (26.0) |
| Other (eg, declined to state, multirace) | 8 (4.0) | 3 (3.0) | 1 (2.0) | 4 (8.0) |
| Education, %† |  |  |  |  |
| High school/GED or less | 19 (9.5) | 11 (11.0) | 4 (8.0) | 4 (8.0) |
| College - 1 year to 3 years | 42 (21.0) | 19 (19.0) | 9 (18.0) | 14 (28.0) |
| College - 4 years or more | 63 (31.5) | 35 (35.0) | 14 (28.0) | 14 (28.0) |
| Post college | 76 (38.0) | 35 (35.0) | 23 (46.0) | 18 (36.0) |
| Annual family income, % |  |  |  |  |
| < \$35,000 | 58 (29.0) | 31 (31.0) | 12 (24.0) | 15 (30.0) |
| \$35,000- <\$55,000 | 37 (18.5) | 19 (19.0) | 8 (16.0) | 10 (20.0) |
| \$55,000- <\$75,000 | 22 (11.0) | 12 (12.0) | 6 (12.0) | 4 (8.0) |
| >=\$75,000 | 83 (41.5) | 38 (38.0) | 24 (48.0) | 21 (42.0) |
| Digital health literacy, %† |  |  |  |  |
| Low 1-1.999 | 0 (0.0) | 0 (0.0) | 0 (0.0) | 0 (0.0) |
| Medium 2-2.999 | 24 (12.0) | 13 (13.0) | 6 (12.0) | 5 (10.0) |
| High 3-4 | 176 (88.0) | 87 (87.0) | 44 (88.0) | 45 (90.0) |
| PHQ-9 category, % |  |  |  |  |
| Minimal depression 0-4 | 4 (2.0) | 2 (2.0) | 2 (4.0) | 0 (0.0) |
| Mild depression 5-9 | 28 (14.0) | 16 (16.0) | 6 (12.0) | 6 (12.0) |
| Moderate depression 10-14 | 116 (58.0) | 56 (56.0) | 28 (56.0) | 32 (64.0) |
| Moderately severe depression 15-19 | 52 (26.0) | 26 (26.0) | 14 (28.0) | 12 (24.0) |
| GAD-7 category, % |  |  |  |  |
| Minimal anxiety 0-4 | 3 (1.5) | 1 (1.0) | 0 (0.0) | 2 (4.0) |
| Mild anxiety 5-9 | 42 (21.0) | 21 (21.0) | 11 (22.0) | 10 (20.0) |
| Moderate anxiety 10-14 | 155 (77.5) | 78 (78.0) | 39 (78.0) | 38 (76.0) |
| PHQ-9 score† | 12.3 ± 3.5 | 12.1 ± 3.6 | 12.2 ± 3.6 | 12.7 ± 3.1 |
| GAD-7 score† | 10.9 ± 2.4 | 10.9 ± 2.4 | 11.0 ± 2.3 | 10.8 ± 2.6 |
| PHQ-9 and GAD-9 inclusion criteria |  |  |  |  |
| PHQ-9 (10-19) only | 45 (22.5) | 22 (22.0) | 11 (22.0) | 12 (24.0) |
| GAD-7 (10-14) only | 32 (16.0) | 18 (18.0) | 8 (16.0) | 6 (12.0) |
| Both | 123 (61.5) | 60 (60.0) | 31 (62.0) | 32 (64.0) |

Abbreviations: GAD-7, Generalized Anxiety Disorder-7; GED, general educational development; HADS, Hospital Anxiety and Depression Scale; PHQ-9, Patient Health Questionnaire-9.
\*Values are mean ± SD unless noted otherwise.
†Randomization co-variates: age, sex, race/ethnicity, education, digital health literacy, PHQ-9, and GAD-7.

All 200 participants completed baseline data; 194 (97.0%) were assessed at 18 weeks and 141 (70.5%) had quality fMRI data at both time points and were included in the primary analysis (Figure 1). Baseline characteristics did not differ significantly between participants with and without quality fMRI data (Supplement, Table S1). Among Lumen-coached participants, 89 (89.0%) completed at least 4 PST sessions (minimum therapeutic dose) and 73 (73.0%) completed all sessions; among human-coached participants, 46 (92.0%) completed at least 4 sessions and 44 (88.0%) completed all sessions.

### Primary outcome

Mean change in right dlPFC activation from baseline to 18 weeks did not differ significantly between either the Lumen- or human-coached arm and waitlist control (Table 2); therefore, the noninferiority analysis between the Lumen- and human-coached arms was not performed.

**Table 2.** Treatment effects on neural and patient-reported mechanistic targets.

|  |  |  |  | Superiority test |  |  |  |  |  | Noninferiority test |  |  |
| --- | --- | --- | --- | --- | --- | --- | --- | --- | --- | --- | --- | --- |
| Neural target | Unadjusted mean ± SD | | | Lumen vs Waitlist | | | Human vs Waitlist | | | Lumen vs Human | | |
|  | Lumen | Human | Waitlist | Mean difference (95% CI)* | P/P<sub>adj</sub>*<sup>‡</sup> | Cohen d<sup>†</sup> (Lumen-Waitlist) | Mean difference (95% CI)* | P/P<sub>adj</sub>*<sup>‡</sup> | Cohen d<sup>†</sup> (Human-Waitlist) |  |  |  |
| Right dlPFC | n=71 | n=36 | n=34 |  |  |  |  |  |  |  |  |  |
| baseline | 0.26 ± 0.49 | 0.30 ± 0.44 | 0.14 ± 0.47 | | | | | | | Not Applicable | | |
| change at 18 weeks | -0.01 ± 0.61 | 0.10 ± 0.77 | -0.03 ± 0.60 | 0.02 (-0.23, 0.27) | .89/.91 | 0.03 | 0.13 (-0.2, 0.46) | .44/.55 | 0.18 | | | |
| Patient-reported target | Unadjusted mean ± SD | | | Lumen versus Waitlist | | | Human versus Waitlist | | | Lumen versus Human | | |
|  | Lumen | Human | Waitlist | Mean difference (95% CI)<sup>‡</sup> | P/P<sub>adj</sub> (2-sided)<sup>‡</sup> | Cohen d<sup>†</sup> (Lumen-Waitlist) | Mean difference (95% CI)<sup>‡</sup> | P/P<sub>adj</sub> (2-sided)<sup>‡</sup> | Cohen d<sup>†</sup> (Human-Waitlist) | Mean difference (90% CI)<sup>‡</sup> | P (1-sided)<sup>‡</sup> | Cohen d<sup>†</sup> (Human-Lumen) |
| SPSI-R:S raw score§ | n=96 | n=48 | n=49 |  |  |  |  |  |  |  |  |  |
| baseline | 12.01 ± 2.91 | 11.08 ± 2.75 | 11.40 ± 2.98 | | | | | | | | | |
| change at 18 weeks | 1.39 ± 2.63 | 1.77 ± 2.53 | 0.52 ± 2.26 | 1.04 (0.23, 1.84) | .01/.03 | 0.34 | 1.11 (0.18, 2.04) | .02/.04 | 0.52 | -0.07 (-0.75, 0.61) | .45 | 0.15 |
| PPO raw score§ | n=96 | n=48 | n=49 |  |  |  |  |  |  |  |  |  |
| baseline | 11.22 ± 3.98 | 10.18 ± 3.78 | 10.06 ± 4.61 | | | | | | | | | |
| change at 18 weeks | 1.91 ± 3.86 | 2.19 ± 4.25 | 0.94 ± 4.27 | 1.46 (0.21, 2.71) | .02/.04 | 0.24 | 1.26 (-0.18, 2.7) | .09/.15 | 0.29 | 0.2 (-0.86, 1.25) | .60 | 0.07 |
| NPO raw score§ | n=96 | n=48 | n=49 |  |  |  |  |  |  |  |  |  |
| baseline | 9.60 ± 4.62 | 10.38 ± 4.05 | 10.94 ± 4.37 | | | | | | | | | |
| change at 18 weeks | -2.32 ± 4.09 | -3.10 ± 4.21 | -1.08 ± 4.06 | -1.71 (-3.02, -0.39) | .01/.03 | 0.30 | -2.25 (-3.77, -0.74) | .004/.02 | 0.49 | 0.54 (-0.56, 1.65) | .28 | 0.19 |
| RPS raw score§ | n=96 | n=48 | n=49 |  |  |  |  |  |  |  |  |  |
| baseline | 11.11 ± 4.40 | 9.90 ± 4.23 | 11.74 ± 4.49 |  |  |  |  |  |  |  |  |  |
| change at 18 weeks | 1.48 ± 4.38 | 1.56 ± 4.32 | 0.04 ± 3.51 | 1.07 (-0.17, 2.31) | .09/.15 | 0.35 | 0.5 (-0.96, 1.95) | .50/.59 | 0.39 | 0.57 (-0.48, 1.63) | .73 | 0.02 |
| ICS raw score§ | n=96 | n=48 | n=49 |  |  |  |  |  |  |  |  |  |
| baseline | 5.69 ± 4.27 | 6.44 ± 4.00 | 5.88 ± 4.01 |  |  |  |  |  |  |  |  |  |
| change at 18 weeks | -0.46 ± 3.98 | -0.40 ± 3.99 | -0.18 ± 4.01 | -0.36 (-1.57, 0.86) | .56/.65 | 0.07 | 0.04 (-1.37, 1.45) | .95/.95 | 0.05 | -0.4 (-1.43, 0.63) | .68 | -0.02 |
| AS raw score§ | n=96 | n=48 | n=49 |  |  |  |  |  |  |  |  |  |
| baseline | 7.01 ± 4.47 | 7.88 ± 4.87 | 7.96 ± 4.42 |  |  |  |  |  |  |  |  |  |
| change at 18 weeks | -0.77 ± 4.10 | -1.58 ± 4.09 | -0.37 ± 4.64 | -0.77 (-2.05, 0.52) | .24/.33 | 0.09 | -1.24 (-2.72, 0.25) | .10/.16 | 0.28 | 0.47 (-0.62, 1.56) | .30 | 0.20 |

Abbreviations: AS, avoidant problem-solving style; CI, confidence interval; FDR, false discovery rate; ICS, impulsive/careless problem-solving style; NPO, negative problem orientation; PPO, positive problem orientation; RPS, rational problem-solving style; SD, standard deviation; SPSI-R:S, Social Problem-solving Inventory-Revised Short Form.
\*Student *t* tests. Both *P* values and FDR-adjusted *P* values were reported.
†Cohen *d* was calculated using the mean difference between 2 groups divided by the pooled SD.
‡Model-based mean differences with 95% CI and *P* and FDR-adjusted *P* values (2-sided) for superiority tests, and with 90% CI and *P* values (1-sided) for noninferiority tests, were generated from analysis of covariance (ANCOVA) models, adjusted for baseline values of the outcome of interest.
§SPSI-R:S score=(PPO raw score/5)+(20- NPO raw score)/5+ (RPS raw score/5)+(20- ICS raw score)/5+(20- AS raw score)/5; the higher the score the better the overall problem-solving ability. Subscales (PPO, NPO, RPS, ICS, and AS) are raw scores without reversal.

### Secondary outcomes

#### Neural mechanistic targets

Secondary neural targets did not differ significantly between either the Lumen- or human-coached arm and waitlist control (Supplement, Table S2).

#### Patient-reported mechanistic targets

Compared with waitlist control, both the Lumen-coached (Cohen d=0.34) and human-coached (Cohen d=0.52) arms showed significantly greater increases in overall problem-solving ability from baseline to 18 weeks. Mean change in overall problem-solving ability did not differ significantly between Lumen- and human-coached participants in either the ITT or per-protocol noninferiority test (Table 2; Supplement, Table S3).

Compared with waitlist control, Lumen-coached participants had significantly greater improvements from baseline to 18 weeks in PPO, NPO, worry, and positive and negative affect (Cohen d range, 0.24-0.5; changes in positive affect no longer significant after FDR correction). Changes in RPS, ICS, AS, and dysfunctional attitudes did not differ significantly between Lumen-coached and waitlist control participants (Table 2; Supplement, Table S2).

Similarly, compared with waitlist control, human-coached participants had significantly greater improvements from baseline to 18 weeks in NPO, worry, and negative affect (Cohen d range, 0.49-0.84). Changes in PPO, RPS, ICS, AS, dysfunctional attitudes, and positive affect did not differ between human-coached and waitlist control participants (Table 2; Supplement, Table S2).

#### Symptom and functional outcomes

Compared with waitlist control, both the Lumen-coached (Cohen d=0.49) and human-coached (Cohen d=0.65) arms showed significantly greater reductions in psychological distress due to depressive and anxiety symptoms from baseline to 18 weeks. Mean change in psychological distress did not differ significantly between Lumen- and human-coached participants in either the ITT or per-protocol noninferiority test (Table 3; Supplement, Table S3).

**Table 3.** Treatment effects on patient-reported symptom outcomes.

| Unadjusted mean ± SD |  |  |  | Superiority test |  |  |  |  |  | Noninferiority test |  |  |
| --- | --- | --- | --- | --- | --- | --- | --- | --- | --- | --- | --- | --- |
|  |  |  |  | Lumen versus Waitlist |  |  | Human versus Waitlist |  |  | Lumen versus Human |  |  |
| Symptom outcome | Lumen | Human | Waitlist | Mean difference (95% CI)* | P/P<sub>adj</sub> (2-sided)* | Cohen d† (Lumen-Waitlist) | Mean difference (95% CI)* | P/P<sub>adj</sub> (2-sided)* | Cohen d† (Human-Waitlist) | Mean difference (90% CI)* | P (1-sided)* | Cohen d† (Human-Lumen) |
| Psychological distress‡ | n=97 | n=48 | n=49 |  |  |  |  |  |  |  |  |  |
| baseline | 18.38 ± 5.61 | 18.98 ± 4.52 | 19.68 ± 5.75 |  |  |  |  |  |  |  |  |  |
| change at 18 weeks | -5.85 ± 6.38 | -7.33 ± 8.04 | -2.76 ± 5.99 | -3.56 (-5.69, -1.43) | .001/.006 | 0.49 | -4.81 (-7.28, -2.35) | <.001/<.001 | 0.65 | 1.26 (-0.54, 3.05) | .23 | 0.21 |
| Depression§ | n=97 | n=48 | n=49 |  |  |  |  |  |  |  |  |  |
| baseline | 7.09 ± 3.32 | 7.84 ± 3.05 | 8.34 ± 3.01 |  |  |  |  |  |  |  |  |  |
| change at 18 weeks | -2.53 ± 3.22 | -3.27 ± 4.47 | -1.92 ± 3.33 | -1.15 (-2.25, -0.04) | .04/.08 | 0.19 | -1.6 (-2.87, -0.33) | .01/.03 | 0.34 | 0.45 (-0.47, 1.38) | .28 | 0.20 |
| Anxiety§ | n=97 | n=48 | n=49 |  |  |  |  |  |  |  |  |  |
| baseline | 11.29 ± 3.59 | 11.14 ± 3.10 | 11.34 ± 3.59 |  |  |  |  |  |  |  |  |  |
| change at 18 weeks | -3.32 ± 4.10 | -4.06 ± 4.49 | -0.84 ± 3.96 | -2.43 (-3.72, -1.14) | <.001/<.001 | 0.61 | -3.22 (-4.72, -1.73) | <.001/<.001 | 0.76 | 0.79 (-0.3, 1.88) | .22 | 0.18 |

Abbreviations: CI, confidence interval; FDR, false discovery rate; HADS, Hospital Anxiety and Depression Scale; SD, standard deviation.
\*Model-based mean differences with 95% CI and P and FDR-adjusted P values (2-sided) for superiority tests, and with 90% CI and P values (1-sided) for noninferiority tests, were generated from analysis of covariance (ANCOVA) models, adjusted for baseline values of the outcome of interest.
†Cohen d was calculated using the mean difference between two groups divided by the pooled standard deviation.
‡The HADS includes 7 questions about anxiety symptoms and 7 questions about depressive symptoms. Each item is scored from 0-3. Total score for the entire scale ranges from 0 to 42, with higher scores indicating more psychological distress due to symptoms of depression and anxiety.
§Scores range from 0-21: normal, 0-7; borderline abnormal, 8-10; abnormal, 11-21.

Compared with waitlist control, Lumen-coached participants had significantly greater reductions from baseline to 18 weeks in depressive and anxiety symptoms (changes in depressive outcomes no longer significant after FDR correction), disability, percent impairment at work, percent overall work productivity loss, and percent activity impairment (Cohen d range, 0.19-0.61) (Table 3; Supplement, Table S4). Change in percent work time missed did not differ significantly between Lumen-coached and waitlist control participants.

Similarly, compared with waitlist control, human-coached participants had significantly greater reductions from baseline to 18 weeks in depressive and anxiety symptoms, disability, percent impairment at work, percent overall work productivity loss, and percent activity impairment (Cohen d range, 0.34-0.76) (Table 3; Supplement, Table S4). Change in percent work time missed did not differ significantly between human-coached and waitlist control participants.

### Associations between change in problem-solving ability and symptoms

Mediation analysis was not performed on the neural targets because the changes did not differ significantly between either intervention arm and waitlist control. Given that overall problem-solving ability improved significantly more in both interventions relative to waitlist control (path A), we conducted an exploratory mediation analysis using it as the mediator, consistent with the theoretical basis of PST.

For path B, improvement in overall problem-solving ability correlated significantly with reductions in psychological distress and anxiety symptoms when comparing changes from baseline to 18 weeks between the Lumen-coached and waitlist control arms and separately between the human-coached and waitlist control arms (Supplement, Table S5).

Change in overall problem-solving ability was not significantly correlated with change in depressive symptoms in either the Lumen-or human-coached arm compared with waitlist control.

### Safety

There were 36 adverse events: 1 serious (expected, unrelated), involving a hospitalization for pneumonia, and 35 nonserious (5 unexpected, unrelated; 30 expected, unrelated) (Supplement, Table S6). There were no deaths.

## Discussion

This phase 2, 3-arm experimental-therapeutics trial evaluated the mechanisms and potential efficacy of Lumen, a rule-based, voice-based AI coach for PST. Among 200 adults with untreated, clinically significant depressive and/or anxiety symptoms, neither Lumen-coached nor human-coached PST resulted in significant changes in the primary neural target compared with waitlist control. However, preliminary findings of a non-neural candidate mechanism and a clinical signal warrant further testing.

This study did not replicate the Lumen phase 1 trial finding of neural target engagement (i.e., right dlPFC) for cognitive control; findings for human-coached PST were also not significant. Despite high overall retention, 30% of participants lacked acceptable fMRI data because of excessive motion or poor signal-to-noise ratios, limiting the validity of neural target analyses. Furthermore, neural correlates of improvements in problem-solving ability and clinical symptoms may extend beyond the prespecified individual ROIs. In another study, we demonstrated that restoration of both activity and functional connectivity in key dorsal prefrontal regions of the cognitive control circuit was associated with improved problem-solving ability and depressive symptoms following human-coached PST.^45^ Importantly, these circuit-level changes persisted over time, with effects observed across five assessment points spanning 24 months. Further studies using similar circuit-level analysis and a data-driven, whole-brain network approach may yield insights regarding neural target engagement for Lumen- and human-coached PST.

Although the secondary findings on Lumen-coached PST’s effects on patient-reported target engagement and clinical and functional outcomes are preliminary, they provide signals for further investigation. As with human-coached PST, Lumen-coached PST significantly improved problem-solving ability—a theory-based behavioral target related to cognitive control—and reduced psychological distress due to depressive and anxiety symptoms, compared with waitlist control. These changes were significantly correlated, suggesting a possible mediating relationship, although the concurrent measurement design precludes causal inference. These findings offer exploratory evidence of a non-neural candidate behavioral mechanism (problem-solving) and clinical signal (psychological distress). Lumen- and human-coached PST also converged on significantly greater improvements, relative to waitlist control, in emotion reactivity (worry and negative affect) and functional impairment (disability, work and activity impairments, and work productivity). This consistent pattern lends support for Lumen-coached PST’s promise as a viable alternative to human-coached PST, though confirmatory trials are warranted.

The noninferiority tests on secondary outcomes were exploratory. In the absence of prespecified noninferiority margins, we benchmarked results against minimal clinically important differences (MCIDs) for the HADS symptom outcomes to interpret their potential clinical significance. Across studies of different populations and methodologies, MCIDs for the HADS range from 2.1 to 8.5 for psychological distress and from 1.1 to 5.6 for the depressive and anxiety subscales.^46–49^ Noninferiority test results showed that the magnitudes of the differences between Lumen- and human-coached PST in the HADS outcomes fell on the low end of these MCID ranges, providing preliminary data for a future noninferiority trial testing Lumen as a clinically acceptable alternative to human-delivered PST.

To our knowledge, Lumen is the first voice-based AI coach for mental health treatment evaluated in randomized clinical trials using an experimental therapeutics framework, with an explicit focus on mechanisms of action in intervention development and testing.^20^ In line with this framework, this study investigated theory-driven neural and patient-reported mechanistic targets. The inclusion of human-coached PST as an active comparator was also a notable strength.^50^ The findings are consistent with recent meta-analyses, which concluded that chatbot interventions modestly reduce depressive and anxiety symptoms;^7,13,51,52^ however, heterogeneous designs, scant mechanistic evidence, and few voice AI intervention studies underscore the need for more rigorous trials.^13^

With further investigation, especially of behavioral mechanisms and clinical symptoms, Lumen has the potential to become a resource for addressing gaps in mental health care. For example, with this digital infrastructure, patients could potentially be “prescribed” Lumen, monitored electronically, and based on progress, transitioned to other care options (e.g., a human psychologist). Lumen’s “device-as-coach” architecture supports scalability, clinical translation, and longitudinal monitoring.^53^ Its rule-based framing also prioritizes safety and treatment fidelity through PST-aligned design, minimizing risks associated with hallucinations and inaccurate responses—key concerns with contemporary AI chatbots.^54^

This study has several limitations. The sample was relatively small and limited to English-speaking individuals with clinically significant but nonsevere depressive and/or anxiety symptoms. The study also excluded some individuals who may have benefited from the intervention because of design and feasibility constraints, including those unable to undergo fMRI, limited internet access, and non-English speakers. In addition, the rate of missing fMRI data was higher than anticipated, reducing the analytic sample and power for the primary neural outcome. Although completion of a minimum therapeutic dose was similar across active arms, full completion was lower for Lumen-coached than for human-coached PST, indicating that future refinement should address sustained engagement across the full intervention course.

Among adults with untreated, clinically significant depression and/or anxiety, Lumen-coached PST did not significantly engage the prespecified neural target for cognitive control, but significantly improved problem-solving ability and psychological distress compared with waitlist control, with modest effects that did not differ statistically from those of human-coached PST. These preliminary findings require confirmation in adequately powered trials.

## Data Availability Statement

Data used in the preparation of this manuscript will be submitted to the National Institute of Mental Health (NIMH) Data Archive (NDA). NDA is a collaborative informatics system created by the National Institutes of Health to provide a national resource to support and accelerate research in mental health. Those wishing to use this data can contact the corresponding author for the dataset identifier and make a request to the NIMH (visit https://nda.nih.gov/). This manuscript reflects the views of the authors and may not reflect the opinions or views of the NIH.

## Code Availability Statement

The SAS code for the statistical analysis is available upon request. Access requests should be directed to the study statistician.

## Author Contributions

Drs. Ma and Xiao had full access to all of the data and took responsibility for the data and accuracy of the data analysis. Drs. Kannampallil and Ajilore contributed equally to this manuscript.

*Concept and design:* Kannampallil, Ajilore, Smyth, Ma.

*Acquisition, analysis, or interpretation of data:* Kannampallil, Ajilore, Smyth, Barve, Ronneberg, Lv, Kumar, Garcia, Aborisade, Wittels, Tang, Chinnakotla, Xiao, Ma.

*Critical review of the manuscript for important intellectual content:* Kannampallil, Ajilore, Smyth, Barve, Ronneberg, Lv, Kumar, Garcia, Aborisade, Wittels, Tang, Chinnakotla, Xiao, Ma.

*Statistical analysis:* Kannampallil, Ajilore, Lv, Xiao, Ma

*Obtained funding:* Kannampallil, Ajilore, Smyth, Ma

*Administrative, technical, or material support:* Kannampallil, Ajilore, Lv, Kumar, Wittels, Xiao, Ma.

*Supervision:* Kannampallil, Ajilore, Ma

