## Supplement for "Neural, Behavioral, and Clinical Outcomes of a Voice-based AI Coach for Depression and Anxiety: A Phase 2 Randomized Trial"

### Table of Contents

### Section A: Institutional Review Board Approved Protocol………………………………………………….……2

Section B: Inclusion and Exclusion Criteria………………………………………………………………..……33

### Section C: Lumen Architecture……………………………………………………………………………….…34

### Section D: Participant Interactions with Lumen ……………………………………………………...…………36

### Section E: Components of PST Implemented in Lumen…………………………………………………...……37

### Section F: Neuroimaging Acquisition and Processing Protocol ………………………………………...………40

### Section G: Treatment Effects on Secondary Neural and Patient-reported Target Measures and Patient-reported Functional Outcomes …………………………………………………………………………………………....43

Section H: Associations between Problem-solving and Clinical Symptom Outcomes (Path B)……..…………50

### Section I: Adverse Events Summary………………………………………………………………………….…51

### Section A: Institutional Review Board Approved Protocol (UIC IRB Protocol #STUDY2022-1052, Version 6, Aproved on April 8, 2024)

**REVISION HISTORY**

| **Revision #** | **Version Date** | **Summary of Changes** | **Consent Change?** |
| --- | --- | --- | --- |
| V2 | 03.30.2023 | - 10.0 Recruitment methods: Recruitment and Screening. Added mail invitation service in Recruitment and screening | No |
| V3 | 05.17.2023 | - 15.0 Risks to Subjects: Updated the “Protection against risks of self-harm” section to clarify that the study psychiatrist will reach out via the phone, email, or text within 3-4 calendar days to participants who report suicidal ideation on PHQ-9, but with no active plan. | No |
| V4 | 07.11.2023 | - 10.0 Recruitment Methods: Updated secondary recruitment strategies to include social media | No |
| V5 | 09.27.2023 | - 11.0: Proecdures Involved: Randomization and blinding: Removed education from the list of randomization covariates as this variable was not used in randomziation. | No |
| V6 | 04.05.2024 | - 10.0 Recruitment Methods: Specified the three UI Health clinics where approved recruitment materials will be distributed. | No |

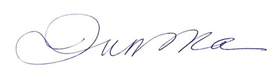
___________________	6/1/2026

PI Signature	Date

### 1.0 Study Summary

Depression and anxiety are the leading causes of disability and lost productivity, and are often underdiagnosed and undertreated owing to access, cost, and stigma barriers. Novel and scalable psychotherapies are urgently needed. Advances in artificial intelligence (AI) offer a transformative opportunity to develop intelligent voice assistants as virtual health agents accessible on personal devices. Meanwhile, major advances in human neuroscience have fueled a paradigm shift to study brain mechanisms underlying behavioral health interventions. Leveraging emerging science in these transdisciplinary areas, this project aims to develop and rigorously test a novel voice-enabled, AI virtual coach named Lumen, trained on Problem Solving Therapy (PST), for patients with mild-to-moderate, untreated depressive and/or anxiety symptoms. The project will investigate the effects of Lumen on these symptoms and on engagement of a priori neural targets—amygdala for emotional reactivity and dorsal lateral prefrontal cortex (dlPFC) for cognitive control—as putative underlying mechanisms. The project has 2 phases: R61 and R33 (see Figure 1). Phase 1 was completed (UIC IRB protocol #2020-0918). Phase 2 will be the sole focus of this protocol and includes a 3-arm randomized clinical trial (RCT) in which 200 newly enrolled participants meeting the same depression and anxiety criteria as in Phase 1 will be randomly assigned in a 2:1:1 ratio to 1 of 3 arms: Lumen Coached PST (n=100), Human Coached PST as active treatment control (n=50), and optional (delayed) Lumen Coached PST (waitlist control; n=50)

**The SPEAC Project**

**Phase** **2: 3-arm RCT** (R33)

**Phase 2 (R33)**

3-arm pilot RCT with 200 participants:

- Lumen Coached PST
- Human Coached PST (active control)
- Optional (delayed) Lumen Coached PST (waitlist control)

**Phase 1 (R61)**

**Pilot RCT**

RCT (3-arm) with 200 participants

**Phase 1:** **Developmental** **Research** (R61)

**Aim 1**

Lumen design, development, and formative evaluation in **2 stages**

- Scenario-based evaluations
- Formative user study

**Aim 2**

2-arm pilot RCT with 60 participants

- Lumen Coached PST
- Optional (delayed) Lumen Coached PST (waitlist control)

**Aim 1**

Lumen design, development, and formative evaluation in **3 stages**

1. Scenario-based evaluations
2. Formative user study

**Aim 2**

Pilot RCT (2-arm) with 60 participants to demonstrate **feasibility** and **acceptability** and test **neural target engagement**

**Figure 1.** SPEAC Project Overview

### 2.0 Objectives

The SPEAC Phase 2 (SPEAC-2) study has 2 specific aims.

**Aim 1: Confirm neural target engagement in a 3-arm RCT**. Primary superiority hypothesis: Compared with waitlist controls, Lumen participants will show *significantly greater* improvements from week 0 to 18 in the neural target, right dlPFC engaged by the go/nogo task, which met the Go criteria for neural target engagement in Phase 1. Secondary noninferiority hypothesis: Compared with human-coached PST participants, Lumen participants will show *no less* improvements (i.e., noninferiority) in the same neural target (relative to the waitlist control) from week 0 to 18. Additionally, we will assess left dlPFC engaged by the go/nogo task and bilateral amygdala engaged by negative facial emotion tasks as pre-specified secondary neural targets. Other neural targets may be identified post-hoc based on whole brain analysis.

**Aim 2: Examine the relationships of neural target engagement to outcomes.** Hypotheses: (1) both Lumen and human-coached PST will lead to better changes in patient-reported outcomes (PROs; depression and anxiety symptoms, psychosocial functioning, quality of life) from week 0 to 18 than waitlist control; and (2) neural target engagement mediates these outcome changes. Additionally, we also hypothesize that both Lumen and human-coached PST will lead to greater improvements in ecological daily assessments of mood, stress and coping over time than waitlist control.

### 3.0 Background

The prevalence of depression in the United States has increased multiple fold to approximately 32% during the COVID-19 pandemic.^1^ Correspondingly, >40 million adults (~19%) have anxiety disorders,^2^ often with co-morbid depressive symptoms. These conditions often go undiagnosed and untreated. Alarmingly, half of Americans with depression and 63% of those with anxiety do not receive any treatment, with the lowest rates among racial/ethnic minorities and persons of low socioeconomic status.^3-6^ People with mental illness often prefer psychotherapy to medication.^7-9^ Yet, the reach and adoption of proven psychotherapies in mental health or general medical settings are limited, owing to barriers such as low reimbursement, provider shortage, patients’ lack of time and transportation, and stigma.^10-13^ As such, there is a critically unmet need for empirically validated psychotherapies that have low cost, avoid stigma, and can be scaled to help address public health and health equity.

Technology-based interventions have quickly grown as a viable option for treatment delivery to address cost, access, and stigma barriers for traditional mental health services. The general public is highly receptive (76% reporting interest) to mental health monitoring and counseling using internet and mobile technologies,^14^ and individuals with mental health concerns often prefer to seek help online rather than in person.^15^ Studies have tested a broad range of technologies for delivering bona fide psychotherapies.^16,17^ The strength of the evidence on early-generation technology interventions led to depression and anxiety treatment guidelines recommending computerized cognitive behavioral therapies using web-based or interactive voice response systems.^18^ Digital mental health interventions have since been evolving with technological advances. Several meta-analyses have documented growing evidence on the utility of mobile interventions for the management of depression and anxiety.^19,20^ One latest area of prolific technological growth in AI is around voice-based personal assistants, which are now nearly ubiquitous in personal home or mobile devices—with recent reports^21,22^ describing that worldwide sales of Alexa devices crossing 100 million and that of Google Home devices crossing 50 million.

With the advances in AI technology, creating persuasive systems that mimic human-like behaviors has become a realistic endeavor.^23,24^ The acceptability of AI-based systems in pragmatic applications is bolstered by studies that have shown that people are more willing to disclose personal information to a virtual agent than when they believe it is human-operated.^25^ Empirical and theoretical research in the field of human-computer interface has shown that people anthropomorphize computers and complex technology^26,27^ and form social attitudes and behaviors and emotional responses towards them. Experiments showed conversational partners whether perceived to be text-based chatbots or humans were equally effective at creating emotional, relational, and psychological benefits,^28^ and establishing strong therapeutic alliances.^29^ Importantly, older adults and people with low reading and/or computer literacy find virtual agents approachable and usable,^30,31^ making these tools particularly relevant for addressing mental health disparities. Much of the development efforts to date have been centered around *text-based* conversational agents,^32^ which have been shown to be feasible in a variety of settings,^33,34^ and for depression and anxiety counseling.^35^ Although promising, text-based intelligent conversational agents have limitations including a lack of personalized interaction and difficulty for use among older adults or people with low literacy.^32,36^ In contrast, AI-powered personal assistants in dedicated voice devices (e.g., Amazon Echo, Google Home) and voice assistant applications (e.g., Alexa, Google Assistant, Siri) have advanced features to engage in human-like conversations. By utilizing automatic speech recognition, natural language processing, and deep learning algorithms, these voice assistants can be transformed from performing routine tasks (e.g., reporting current weather) to more sophisticated and context-specific health intervention tools. Research to develop and test such interventions is in its infancy.

The recently completed SPEAC-1 study was designed to develop and pilot test the effect of a voice-enabled, AI virtual coach named Lumen on engaging empirically supported neural targets as well as the relationship of change in neural targets to change in depression and anxiety symptoms among 63 adults with mild-to-moderate depression and/or anxiety. The findings showed that the Lumen intervention resulted in medium effects (Cohen’s d=0.49 to 0.55) for depression and anxiety symptom reductions, compared with a waitlist control condition. The intervention effect relative to control met prespecified threshold (d=0.3) for change in activation of the right dlPFC, which correlated with self-reported measures of problem solving (r≥0.4). This voice-based virtual coach delivering PST has showed promising effects on depression and anxiety symptoms and neural and self-reported measures of cognitive control. Accordingly, the SPEAC-2 trial will further rigorously test Lumen against both a waitlist control and an active treatment control condition that will provide human coached PST in a larger study sample.

### 4.0 Study Endpoints

### The study endpoint is at 18 weeks post randomization. See Appendix 1 for schedule of measures and Appendix 2 for description of outcome measures.

### 5.0 Study Intervention

**Lumen Coached PST Intervention.** Lumen is a voice-based virtual coach developed on Amazon’s Alexa platform. Lumen delivers an evidence-based PST program^37,38^ consisting of 8 sessions (4 weekly, followed by 4 biweekly sessions) for patients with mild-to-moderate depression and/or anxiety. PST is patient-driven, where the coach acts as a guide to identify a problem, set a goal, brainstorm solutions, choose a solution, develop an action plan, and to implement and evaluate the plan.^39^ This stepwise approach makes PST appropriate for therapy delivery using a voice-based virtual coach.

Lumen was designed through an iterative user-centered process that involved software developers, interaction designers, psychiatrists, PST experts, and behavioral scientists. Several iterations of the prototype were internally tested; a fully functional prototype underwent feasibility and usability testing with 26 users.^40^ Design was driven by 2 key principles: (a) aligning participants’ voice-based interaction with Lumen similar to the cognitive processes of human communicative interactions,^41^ and (b) configuring the content of the interactions with the principles and process of evidence-based PST.

For participants, Lumen was integrated within the Alexa app on an iPad provided by the study. For each session, participants initiate Lumen PST through the Alexa app with a “Launch Lumen Coach” voice instruction and then complete the assigned PST session. A typical Lumen session lasted ~12 minutes. Participants also complete the PHQ-9 (Patient Health Questionnaire – 9) and GAD-7 (Generalized Anxiety Disorder) before all PST sessions. Upon completion of the entire intervention participants will receive a certificate of completion.

**Lumen Intervention Orientation**. Participants in the Lumen intervention arm attend a Lumen orientation visit (60 minutes) during which they will be given a Lumen intervention tutorial and receive a study iPad, configured to limit access to only the Lumen intervention enabled on the device. Before leaving, participants are scheduled for the first PST session with Coach Lumen. At the end of each PST session, participants are prompted to schedule their next session with Coach Lumen. They receive automated reminder notifications 1 day prior to their next session, on the session day, and have the opportunity to make up missed sessions. As needed, Lumen intervention orientations can be done remotely.

**Human Coached PST Intervention.** Participants in the human coached PST arm will serve as active treatment controls given the demonstrated efficacy of human coached PST in depression and anxiety. They will complete 8 PST sessions with a human coach; first session in-person and remaining sessions via videoconferencing (i.e., Zoom preferred; with phone calls allowed) on the same 4 weekly and then 4 biweekly schedule as that for Lumen participants. Participants will receive a study iPad to use for their PST sessions via zoom. Participants also complete the PHQ-9 and GAD-7 during each session. Participants will receive a certificate of completion.

**Intervention Fidelity Assurance.** All PST sessions that participants complete with Lumen or human coach are audiorecorded. For Lumen, session recordings and transcripts are stored on the Amazon Web Services (AWS) in study configured accounts on a UIC secure server, with coded identifiers. Audio recordings of human-coached PST sessions (recorded via zoom recordings) will be placed on a UIC secure server with coded identifiers as well. Recordings will be used to evaluate treatment fidelity and user-Lumen interaction.

**Optional (delayed) Lumen Coached PST (Waitlist control)**. Waitlist control participants will only complete assessments during the 18-week trial period. At the end of their 18-week assessment, they will have the option to complete 8 PST sessions on their assigned iPads.

### 6.0 Study Timelines

**Figure 2.** Project Timeline & Work Plan (in calendar years and quarters)

|  | 2022-23 | | | | 2023-24 | | | | 2024-25 | | | |
| --- | --- | --- | --- | --- | --- | --- | --- | --- | --- | --- | --- | --- |
|  | Jul | Oct | Jan | Apr | Jul | Oct | Jan | Apr | Jul | Oct | Jan | Apr |
| (Sub-)contractual agreements |  |  |  |  |  |  |  |  |  |  |  |  |
| Refinements of Lumen and MOP and study databases |  |  |  |  |  |  |  |  |  |  |  |  |
| IRB approval of final protocol and procedures |  |  |  |  |  |  |  |  |  |  |  |  |
| Participant recruitment (N=200) |  |  |  |  |  |  |  |  |  |  |  |  |
| Data collection and intervention delivery |  |  |  |  |  |  |  |  |  |  |  |  |
| Data management and QC |  |  |  |  |  |  |  |  |  |  |  |  |
| Data analysis, publications, and data sharing |  |  |  |  |  |  |  |  |  |  |  |  |

Recruitment begins January 2023, and we estimate that it will take 19 months to enroll and randomize 200 eligible participants. Participants will be randomized to the Lumen Coached Group, Human Coached Group, or the Optional (delayed) Lumen Coached Group and followed for 18 weeks. Final data collection will be completed by January 2025 with the remaining of the final year of the project period allocated to final data cleaning and lock, analyses, publications, and trial closeout. The timeline for enrollment and data analysis is as follows:

| January 2023 | Open for enrollment |
| --- | --- |
| August 2023 | Cumulative 25% of target enrolled reached |
| December 2023 | Cumulative 50% of target enrolled reached |
| April 2024 | Cumulative 75% of target enrolled reached |
| August 2024 | Cumulative 100 % of the target enrolled reached |
| January 2025 | Completion of final data collection |
| May 2025 | Submission of primary manuscript |
| June 2025 | Preparation of de-identified data and relevant documentation for data sharing |

**Participant timeline**. Individual participants’ 18-week participation timeline is detailed in Section 11 procedures, and in the study event windows listed in Appendix 3

### 7.0 Inclusion and Exclusion Criteria

Racially/ethnically and socioeconomically diverse participants will be screened for eligibility by trained staff, using a multistep enrollment process (detailed in Section 10) according to the following criteria:

**7.1 Inclusion Criteria**

- Age: ≥ 18 years
- Emotional distress defined by elevated depressive (PHQ-9 scores 10-19) and/or anxious symptoms (GAD-7 scores 10-14)
- Willing and able to provide informed eConsent and HIPAA authorization

**7.2 Exclusion Criteria**

- Unable to speak, read, or understand English for informed consent
- Current pharmacotherapy or psychotherapy (individual or professionally led group therapy) for depression or anxiety (note: participants are not withdrawn post-randomization if they begin pharmacotherapy drugs or start psychotherapy during the study.)
- Suicidal ideation per PHQ-9 with active plan
- Bipolar or psychotic disorder, or current psychiatric treatment
- Weight ≥325 pounds due to brain scanner constraints, MRI contraindications, traumatic brain injuries, and tumor or any other known structural abnormality in the brain
- Severe medical condition (e.g., myocardial infarction or stroke or new cancer diagnosis in the past 6 months, end-stage organ failure, terminal illness) or residence in a long-term care facility
- Diagnosis of cancer (other than non-melanoma skin cancer) that is/was active or treated with radiation or chemotherapy within the past year
- Active alcohol or substance use disorder (including prescription drugs) based on the CAGE Questionnaire Adapted to Include Drugs (CAGE-AID)
- Cognitive impairment based on the Callahan 6-item screener
- Current or planned pregnancy or lactating (<6 months postpartum)
- Participation in other investigational treatment studies that would significantly affect participation in this study, raise safety concerns, and/or confound outcomes (participant may be asked to provide the informed consent of the other study for final decision on exclusion by a study psychiatrist)
- Family/household member of an already enrolled participant or of a study team member
- Plan to move out of the Chicago area during the study period
- Does not have reliable Wi-Fi Internet at home
- Unwillingness to use personal mobile device to receive study text messages
- Investigator discretion for clinical safety or protocol adherence reasons

We will exclude each of the following special populations:

- Adults unable to consent
- Individuals who are not yet adults (infants, children, teenagers)
- Pregnant women
- Prisoners

### 8.0 Vulnerable Populations

Children are excluded from the study as the target age group is 18 years and older. Psychotherapies of depression and anxiety and related risk protections (including protection of patient privacy and confidentiality) for persons under the age of 18 years differ from those for adults. Adults are the focus in this stage of research to develop a neural mechanism-validated, voice-based virtual coach for PST. In addition, participants unable to speak, read and understand English for informed consent are excluded from the study as the research instruments and intervention are administered in English only.

### 9.0 Number of Subjects

Participants with mild-to-moderate, untreated depression and/or anxiety (N=200) will be randomized in a 2:1:1 ratio to 1 of 3 arms: Lumen Coached PST (n=100), Human Coached PST as active control (n=50), and optional (delayed) Lumen Coached PST as waitlist control (n=50). Approximately 3500 subjects may need be enrolled by consenting to screen for this research study. Of these, 200 participants who are determined fully eligible will be randomized into the study.

### 10.0 Recruitment Methods

**Multistep Screening and enrollment process:** To enable efficient recruitment during the trial, we will implement a multi-step, eligibility screening and enrollment process.

**Prescreening.** The primary recruitment strategy utilizes UI Health’s EHR database to identify patients meeting the basic prescreening criteria (e.g., age, absence of exclusionary medical or psychiatric comorbidities, etc.)

The secondary recruitment strategy uses in-clinic referrals or passive recruitment using advertising brochures and flyers at the UI Health outpatient clinics, community clinics and worksites, social media postings (Instagram, Facebook, Reddit). Online postings include a hyperlink or QR code to the study survey in REDCap. Specifically, within UI Health clinics, efforts are focused on reaching out to Primary Care Plus (Suite 3AA), General Internal Medicine (Suite 3A), and Primary Care Adults and Children (Suite 3). Study brochures are distributed in clinic, and providers may refer patients during routine office visits and direct interested patients to the study website or telephone/text line for more information and screening. In addition, UIC listservs are used to distribute recruitment announcements to UIC employees. The UIHealthResearch registry and the New Normal (TNN) registry offered by UIC Center for Clinical and Translational Science are used to distribute recruitment information to potential participants. Based on previous experience using these recruitment strategies, it is anticipated that EHR-based recruitment will be the predominant source, accounting for >56% of randomized participants. Reviews of patient EHR may be performed to screen for diagnostic exclusion criteria. This step is permissible under a waiver of informed consent. A waiver of informed consent and HIPAA preparatory to research are being requested for this process given that medical record screening is minimal risk, will not affect patients’ rights or welfare, and that without it, subject recruitment within the time and funding limits of the study would be impracticable.

**Recruitment and Screening.** Recruitment invitations are sent to patients by email, if an email address is available in EHR, or by texting a link to the “open for recruitment” announcement on the study website. Patients also receive a study invitation via postal mail, using a third party mailing service. The recruitment email, mail, and online announcement describe the study in lay language and contain a secure web link or a QR code to REDCap where patients may give online consent to screen for eligibility, if interested, or decline further contact if they choose to opt out. Patients may also opt out by directly replying to the invitation email or text or by calling the study recruitment phone line. Patients who do not self-screen or opt-out within 2 weeks of the recruitment invitation date will be contacted via phone to assess interest, either by a Twilio-enabled interactive voice response (IVR) message or by a trained study coordinator. Using the Twilio IVR, an automated message from the principal investigator that follows an abbreviated version of the recruitment invitation script for study coordinators offers a brief introduction of the SPEAC-2 study, and options to 1) decline further contact, 2) request a text message with a link to the study website, or 3) request a call back from study staff. Study coordinators will follow up with participants who request a call back or who do not respond. A participant tracking database will be developed in REDCap, which supports Microsoft Excel exports for analysis and reporting on enrollment and screen failures at each step. During initial eligibility screening, participants who score 10 or above on the GAD-7 and/or PHQ-9 survey and who are interested in UI Health’s and other emotional health services will receive a PDF document with this information via email, regardless of their eligibility status.

**Teleorientation with eConsent**. Screened eligible participants are invited to schedule a teleorientation session with a study coordinator via their choice of telephone or online video conference. Teleorientation begins with the coordinator providing the REDCap link to the online consent document and leading the informed consent discussion, as outlined in Section 24, including study procedures, risks and benefits, the voluntary nature of the research, and the pros and cons of being randomly assigned to Lumen Coached, Human Coached or optional (delayed) Lumen Coached Group. Willing participants document their consent electronically using the secure REDCap eConsent framework. Section 24 details the process for obtaining a PDF copy of the signed consent. The study copy is automatically archived in the REDCap file repository. After obtaining the eConsent, the coordinator will complete the final eligibility interview questions (see Appendix 1 screening measures).

### 11.0 Procedures Involved

**Scheduling study procedures.** After teleorientation, participants are scheduled for a baseline assessment visit. During the baseline visit, a trained study coordinator performs fMRI screening and data acquisition and collects baseline data per standardized measurement protocols. Females of child-bearing age who indicate any possibility that they may be pregnant must take a point-of-care urine pregnancy test. At the end of the baseline visit participants receive instructions to complete the initial ecological daily assessment for the next 7 days. Following successful completion of the baseline measures, fully eligible, consented participants are randomized to either Lumen Coached Group, Human Coached Group or optional (delayed) Lumen Coached Group at a 2: 1: 1 allocation.

Participants assigned to the Lumen Coached Group will complete a Lumen orientation in person and 8 PST sessions on an iPad provided by the study. Participants in the Human Coached Group will complete the first PST session in person and 7 remaining PST sessions via videoconferencing with a health coach (i.e., Zoom preferred; with phone calls allowed). They will receive a study iPad to use for their PST sessions via zoom. Section 5 describes the specifics of the intervention. Participants in the optional (delayed) Lumen Coached Group have the option of completing the Lumen orientation and 8 PST sessions on their assigned iPad after their 18-week assessments.

**Randomization and blinding**. A designated staff person performs randomization using a validated online system.^42^ The system implements Pocock and Simon’s minimization, a covariate-adaptive method,^43^ to achieve better-than-chance marginal balance between study arms across multiple key baseline characteristics. Minimization accommodates a greater number of balancing covariates than does stratified randomization.^44^ The system’s computational algorithm automatically adjusts the randomization probability based on the characteristics of all the previously randomized participants, thus minimizing the total covariate imbalance between arms after each new participant is randomized. In this study, randomization covariates include sex, age, race/ethnicity, digital health literacy,^45^ PHQ-9, and GAD-7. Note that, aside from eligibility screening, PHQ-9 and GAD-7 are also process measures used to monitor treatment progress throughout PST sessions. Their inclusion among randomization covariates helps avoid accidental bias at baseline. The online system also applies Efron’s biased-coin method^46^ to protect allocation concealment with the use of nonextreme randomization probabilities. By design, treatment assignments are identifiable to participants and health coaches delivering the intervention. However, blinding of outcome assessment, event adjudication, and data analysis will be enforced. To accomplish the 2:1:1 allocation, participants are randomized into 4 sets, and then 2 of these sets will be combined to form the Lumen Coached Group. The remaining 2 sets form the Human Coached Group and the optional (delayed) Lumen Coached Group, respectively. This method both protects blinding and preserves the allocation ratio at every allocation as per Kuznetsova and Tymofyeyev.^47^ The same method was used in the completed ENGAGE-2^48^ and SPEAC-1 study.

**Assessments**. All participants complete 2 study assessment visits, at baseline (week 0) and at 18 weeks post randomization, and self-complete ecological daily assessments for 7 days every 2 weeks (on weeks 2, 4, 6, 8, 10, 12). The fMRI scan visits are conducted at the 3T MR Research Program located at the UIMC Advanced Imaging Center, a BioRAFT registered facility, and SPEAC study staff work in compliance with all COVID-19 safety guidelines employed by facility management. When scheduling assessment visits, study coordinators instruct participants in the current COVID-19 safety precautions prior to and at the visits.

Assessments (120 minutes)

- fMRI assesses neural target engagement and treatment outcomes. fMRI is a standard technique for measuring and mapping brain activity that is noninvasive and safe. In this study, it is being used simply to gather neuro imaging data, and not to test the safety/efficacy of a device or software. Afterwards, the patient completes blood pressure, height, and weight measurements. (90 minutes)
- Self-report surveys of PST theory-based constructs of emotion (affect, worry) and cognition (problem solving, dysfunctional attitudes) as well as patient outcomes (e.g., depressive and anxiety symptoms, functioning, quality of life). Participants may complete these surveys during or outside their fMRI visits. (30 minutes)

Self-completed Ecological Daily Assessments (<3 minutes each day)

Using the Lumen companion application, participants complete ecological end-of-day assessments of mood, stress, appraisal, and coping for 7 days at weeks 0 and 18, and every 2 weeks (on weeks 2, 4, 6, 8, 10, 12).

### 12.0 Data and Specimen Banking

We will partner with the NIMH and use all NIMH data preparation and sharing policies as a guide to ensure the coded data and results, along with all associated documentation, are submitted to the NIMH Data Archive (NDA), to be specifically deposited to the National Database for Clinical Trials Related to Mental Illness (NDCT). We will ensure we have permission from our participants to disclose coded participant-level data collected as part of this study to researchers who meet all the NDCT requirements for requesting use of the dataset. We will strictly comply with the HIPAA and IRB regulation requirements regarding research use of PHI and appropriate safeguards for sharing coded data.

### 13.0 Sharing of Results with Subjects*

At participant’s request, a brain image from the participant's MRI scan will be emailed to the participant as a PDF attachment. However, the MRI scan data collected in this study are specific to the research and are not optimized to find medical abnormality. The investigators are not responsible for failure to find existing abnormalities with these MRI scans. In the unlikely event that a participant’s MRI scans show a possible abnormality, the study doctor will contact them and, at the participant’s request, refer for a medical follow up for the problem, including a referral to a Primary Care Provider (PCP). If they already have a PCP, the study doctor will acquire permission from them to speak with their PCP and notify them of the results of the MRI scan in order to assist them get the appropriate follow-up. The decision as to whether to proceed with further examination and/or treatment lies solely with the participant and their PCP. The study will not cover the costs of any follow-up actions.

### 14.0 Withdrawal of Subjects

Participants may be withdrawn from the research without their consent under the following circumstances:

- the investigator decides that continuation can be harmful to the participant,
- participant develops a new condition or undergoes treatment that will make their subsequent follow-up data in the study unusable,
- the study is cancelled, or there are other administrative reasons.

In the event a participant withdraws or is asked to leave the study, they will still be compensated for the study activities they have completed.

### 15.0 Risks to Subjects

This is a minimal risk study with prudent measures in place to protect the health and well-being of research participants and their privacy and confidentiality. Risks associated with participation in this study may include the potential for the following:

- Potential for self-harm
- MRI-related injury, discomfort, and distress
- Inconvenience due to study visits
- Patient privacy and confidentiality breach
- Accidental recording.

These risks are largely associated with the characteristics of the patient population to be studied and the procedures involved in the research. The target population includes patients with mild-to-moderate, untreated depressive and/or anxiety symptoms. The risks are reasonable in relation to the anticipated benefits and are minimized by using procedures that are consistent with sound research designs and established research and clinical protocols. The following measures are implemented to minimize potential risks to participants in the study.

**Protection against risks of self-harm**. Some of the questions about depression, thoughts of death and other psychological symptoms and conditions as a part of study assessments may cause discomfort for some participants. However, in general the questions are not particularly intrusive or distressing, and stress is likely transient. In addition, participants are free to refuse to answer any questions. It is widely accepted that asking questions about thoughts of death or suicide does not lead to increased risk of suicide. Nevertheless, in the event that a patient is identified as being suicidal during the screening or follow-up phase of the study, the following self-harm protection protocol (adapted from the UIC IRB approved SPEAC-1 study #2020-0918, is in place to immediately alert the study supervising psychiatrist to assess the patient’s suicidal thoughts by telephone, followed by notification of the participant’s PCP and appropriate clinical action if necessary. If a participant responds “1” (“several days”), “2” (“more than half the days”), or “3” (“nearly every day”) to the PHQ-9 question “Over the last 2 weeks, how often have you been bothered by thoughts that you would be better off dead or thoughts of hurting yourself in some way?”, we further assess the participant’s level of risk by asking “*Do you have a plan for how you would commit suicide?*” and then follow the protocol based on the assessed level of risk. Individuals reporting suicidal ideation on PHQ-9 with an active plan at eligibility screening are excluded from participation. Nonetheless, the risk of emergent suicidality still exists after enrollment. Risk for suicide may be detected when a participant completes the PHQ-9 at the beginning of each PST session. Participants who score 1-3 (see above) on item 9 of the PHQ-9, will be offered the option to call an emergency contact, the National Suicide Prevention Lifeline at 988 or 911 at the time of detection. This information is a pre-programmed script for Lumen. The supervising study psychiatrist, Dr. Ajilore, the intervention manager, Mrs. Ronneberg, receive immediate notifications, as outlined in the table below. The study psychiatrist calls the participant within 3-4 calendar days to conduct an assessment, assess need for further referral, and discuss referral options depending on urgency, needs and preferences, available supports in place, insurance status and routine source of care. The study psychiatrist may refer the participant for immediate comprehensive evaluation (e.g., at a local emergency department), ambulatory psychiatric services, or community resources, with personal accompaniment depending on the evaluation outcome.

| **[If PHQ-9 completed with SPEAC-2 study staff by PHONE]** | |
| --- | --- |
| **If YES, participant has active plan or DECLINE TO STATE for self-harm:**   - Explain to participant: I am concerned for your safety and therefore need to call for help right now. - Get participant's location - Stay on the phone with participant and use another phone to call 911.   [Script: "*I want to report a self-harm alert for a UI Health research participant who has just endorsed suicidal ideation to me - I want to provide his/her name, location, and phone number (any relevant detail the pt provided.)"]*   - Send High-priority Page to on-call Study Physician.   **Research Staff Note**: You do NOT need participant's consent to call 911 if you feel there is a possibility of immediate risk of harm to self or others. | **If NO active plan explain to participant:**  I am not a clinician; however, our study has clinicians who speak with any participant who tells us they've been feeling this way recently. I will have a study doctor contact you within the next 3-4 calendar days. I would also like to give you 2 national helpline and 1 text line numbers that you may find helpful. All numbers are available 24 hours/7 days a week. We will work together to get you feeling better.  National Hopeline Network: 988 Suicide and Crisis Lifeline  National Suicide Prevention Lifeline: 988 Crisis Text line: text HOME to 741741 |
| **[If PHQ-9 completed by Participant ONLINE ]**  [The following pop-up message appears if a participant responds “1” (“several days”), “2” (“more than half the days”), or “3” (“nearly every day”) to the 9th question, regardless of active action plan or not.]  Please note: We do not monitor this screener in real time, if this is an emergency call 911.  For more immediate attention, because you have been bothered by thoughts that you would be better off dead or of hurting yourself in some way in the last 2 weeks, you should call your physician or other healthcare professional right away or go to the emergency room.  You may also call the National Suicide Prevention Lifeline at 988 hotline. You may also text HOME to Crisis Text Line number 741741. All these numbers are available 24 hours every day.  We will have a study doctor contact you within 3-4 calendar days, In the meantime, do not delay seeking medical attention. | |
| **If PHQ-9 completed with SPEAC-2 Health Coach during Intervention Session** | |
| **If YES, participant has active plan for self-harm or DECLINE TO STATE:**   - Explain to participant: I am concerned for your safety and therefore need to call for help right now. - Get participant's location - Stay with participant and call 911 (use another phone if the session is being conducted by phone).   [Script: "*I want to report a self-harm alert for a UI Health research participant who has just endorsed suicidal ideation to me - I want to provide his/her name, location, and phone number (any relevant detail the pt provided.)"]*   - Send High-priority Page to on-call Study Physician.   Research Staff **Note**: You do NOT need participant's consent to call 911 if you feel there is a possibility of immediate risk of harm to self or others. | **If NO active plan, explain to participant:**  I am not a clinician; however, our study has clinicians who speak with any participant who tells us they've been feeling this way recently. I will have a study doctor contact you within the next 3-4 calendar days. I would also like to give you 2 national helpline and 1 text line numbers that you may find helpful. All numbers are available 24 hours/7 days a week. We will work together to get you feeling better.  National Hopeline Network: 988 Suicide and Crisis Lifeline  National Suicide Prevention Lifeline: 988 Crisis Text line: text HOME to 741741 |

**Protection against injury, discomfort, and distress with brain imaging**.

MRI is non-invasive, widely used, and safe. Routine contraindications to MRI include presence of any metal implants (pacemaker, aneurysm clips, neurostimulators, cochlear, eye implants, old or very fresh tattoos). Participants are thoroughly assessed for MRI eligibility during screening using the well-established procedures as in the SPEAC-1 study #2020-0918 A small number of people may feel claustrophobic inside the MRI machine. The study can be immediately stopped via button press if this occurs. Sometimes subjects report a temporary, slight dizziness or light-headedness when they come out of the scanner. Study personnel and technicians are on site during all acquisitions to address any such discomfort. To minimize fatigue, sufficient breaks are provided to participants during the scanning procedure, and the study coordinator conducting the scan regularly inquires as to whether there is anything that s/he can do to facilitate the participant’s comfort. In the event of adverse effects related to MRI scanning, study personnel and medical staff are on-site for consultation and assistance. The UI Hospital Emergency Room is less than a 5-minute drive from the scanning center. If there are any adverse events at any time during the MRI procedure, the study coordinator terminates the scanning session, provides a debriefing, and contacts study psychiatrist (Dr. Ajilore) for assistance and follow-up for the participant.

According to the National Institute of Mental Health (NIMH) Council Workgroup on MRI Research and Practices (September 2005), “there is no known risk of MR brain scanning of a pregnant woman to the developing fetus for scanning at 4T or less, and no known mechanism of potential risks under normal operating procedures.” Notwithstanding, subjects are warned about potential risks not yet discovered in the informed consent form. Before each scan, female participants are asked if they are or are trying to become pregnant and when they had their last menstrual period. Any woman who indicates that she is pregnant will not be scanned. Any woman who indicates that she is trying to become pregnant or who is experiencing a late menstrual period is asked to complete a urine pregnancy test, which if positive will preclude the subject from being scanned.

If study personnel observe any unusual features in the MRI scan at the time of acquisition or an incidental finding, or abnormality on MRI scans, staff requests a clinical neuroradiological report on the scan from the neuroradiologists at the Center for MR Research who are on call to provide these reports as required, as part of the infrastructure of the Center’s facility. During the consenting process, all participants are informed about the potential risks of discovering an incidental finding or abnormality on their MRI scan. If an abnormality is found in a participant’s MRI scan, MPI Dr. Ajilore contacts the participant and refers him/her for medical follow-up for the problem if the participant requests, including a referral to a PCP. If a participant has a PCP, Dr. Ajilore contacts the PCP, at the request of, and with verbal permission from the participant, to inform him/her of the finding on the MRI scan and to help him/her get the participant appropriate follow-up. The decision as to whether to proceed with further examination and/or treatment lies solely with the participant and his/her PCP.

**Inconvenience due to study visits**. In-person data collection visits are required for all participants in order to obtain physical measurements and MRI scans. Although some degree of inconvenience will be associated with these contacts, it will not be excessive.

**Protection against breaches of participant privacy and confidentiality**. All investigators and their staff are adequately trained to protect participant privacy and sign an agreement to do so. All information obtained from research participants during the study are considered strictly confidential and only used and disclosed as permitted under the HIPAA regulations. All eligible participants must sign a HIPAA authorization as part of the informed consent form in order to participate. They also have the option to agree to sharing of their de-identified data through the NIMH Data Archive (NDA). Only aggregate data will be included in scientific presentations and publications resulting from this study.

In addition, the following measures will be taken during the conduct of the study to ensure adequate protection of participant privacy and confidentiality:

- All investigators and staff will be adequately trained to protect participant privacy and sign an agreement to do so.
- Personal identifiable information (PII) and protected health information (PHI) collected for the purpose of the research study will be assigned a unique anonymous study ID number. The study IDs will be used on all study forms and for data storage, tracking and reporting.
- Both the anonymized health information and the information linking the study ID numbers to participants’ identities will be stored in a password protected database on a secure network fitted for protection of such information, which will be accessible only to the research personnel who have a need to know.
- The information linking the study ID numbers to participants’ identities will be stored separately from the anonymized health information.
- We will follow the latest industry standards for data encryption, server authentication, and client authentication to ensure secure data transmissions at all times.
- No participant data, even if de-identified, shall be saved onto any portable devices, such as laptop computers or USB drives.

In addition to the protection measures described above, randomized participants who begin Lumen coached or human coached intervention per group assignment, receive a study iPad which utilizes the in-built encryption and 6-digit passcode protection and is configured with their first name (as per informed consent). Only first name will be used by Coach Lumen or human coach during intervention to facilitate participant engagement for treatment alliance. Additionally, the study iPads are in a “lock down” mode where participants cannot use them for any purposes but for tasks associated with the study.

No study data resides locally on the iPad. In order to maintain anonymity of the intervention participants, no personal identifiers (other than first name) are used during Lumen PST session data acquisition or storage. All Lumen PST session recordings and transcripts acquired by the Amazon Alexa application using the study iPad are stored on the AWS cloud within study configured accounts. The study-established AWS accounts are assigned coded “AWS IDs” (which are paired with “Study Participant ID”) and are accessible only by study staff with a need to know. In case of a potential loss or theft of the study iPad, participants are instructed to immediately notify study staff and the AWS ID account access will be disabled. Recordings of human coached PST sessions are stored on UIC secure server, coded with study IDs.

The SPEAC-2 study is a registered HIPAA account under a standardized Business Associate Addendum (BAA) held by UIC/UI Health with (AWS) which enables the covered entities to be HIPAA compliant.

Finally, Lumen Coached or Human Coached Group participants are able to keep their study iPad for personal use after their AWS ID account access is disabled. Participants will be provided with instructions to remove the Lumen skill (if assigned to this group) and to unlock the iPad to have a fully enabled device to keep. Alternatively, participants may choose to redeem the iPad for $100. Participants in the optional (delayed) Lumen Coached Group may choose to either receive $100 or attend a Lumen Orientation visit to receive training and a Lumen PST-enabled iPad after their 18-week assessments. These options are explained to study participants during the informed consent and reiterated to waitlist control participants who choose to receive Lumen PST after the end of the study.

**Protection against accidental recording**. Accidental recording is a commonly raised concern regarding voice-enabled technologies. In the case of Lumen, the application is installed on the locked-down and encrypted study iPad. In order to prevent accidental recording, the following protections are implemented: First, the iPad has a keypad lock. Recording takes place only in the unlocked mode, after the participant opens the Alexa app, and gives permission verbally (using specific “wake phrases” such as “Launch Lumen Session,”) or by tapping the option on the iPad screen. Second, participants are informed that all Lumen sessions are recorded and give explicit permission for this as part of their informed consent process. Third, if the study iPad is left unattended during an active session, the iPad locks after 10-seconds, and after 30 seconds of no verbal input from the participant, the Lumen session shuts down (preventing any further recording). Once the participant returns, they must go through the process of unlocking iPad and waking Lumen to actively resume their session.

### 16.0 Potential Benefits to Subjects

Eligible participants in the study have mild-or-moderate depressive and/or anxiety symptoms that are not treated. Lumen intervention participants receive PST by interacting with Lumen on a secure study iPad. Waitlist controls have the option to receive Lumen after their 18-week assessments. Active treatment controls will receive the PST sessions with a health coach. PST is a brief skill-enhancing psychotherapy with robust evidence for efficacy in treating depression and anxiety. PST focuses on improving one’s skills involved in solving personal real-life problems and is easy for most people to understand. Its stepwise, patient-driven approach is also easy to follow. Lumen is a voice-based virtual coach that has undergone iterative design development and refinement and rigorous pilot testing. Through interacting with Lumen, participants in the study may benefit from having PST at their fingertips and experience improvements in psychological symptoms, functioning and quality of life. Lumen may downstream carry the potential to be scaled for use by people with depression and/or anxiety who cannot or will not seek professional help or who desire as-needed, personal help beyond what conventional treatment models can offer. Notwithstanding, we cannot guarantee any individual participants will benefit from participating in the study.

### 17.0 Data Management and Confidentiality

**Data collection**. The study coordinators are trained on data collection according to standardized protocols, and their performance is continuously monitored to ensure data quality. The following types of data are collected.

1. Data on diagnoses, prescriptions, clinical encounters, and hospitalizations are abstracted from EHR for eligibility screening and baseline characterization.
2. fMRI data and physical measurements are collected at the UIMC Advanced Imaging Center.
3. Survey data and ecological daily assessments are collected using self- and interviewer-administered questionnaires on REDCap, a HIPAA-compliant server.
4. With participant’s explicit permission, PST sessions are audiorecorded and are stored in study configured accounts on the AWS cloud or UIC secure server, accessible only by study staff.

See Appendix 1 for Schedule of Measures.

**Neuroimaging data management.** There are Linux machines dedicated to various aspects of image transfer and storage available to the MPI Dr. Ajilore and team within the UIC Psychiatric Institutes. The Institute also houses an imaging processing server for data storage and analysis (Dell PowerEdge R900, 2.13 GHz, Xenon Four Cores, 128GB RAM with 6 TB of data storage). These machines are networked with a T3 line to facilitate image transfer and back-up of data from the Center for MR Research. The T3 line is connected to the University network and firewall shielded from the outside. These machines are for the sole use of the research team and as such the research team has its own log in for the system with direct access from our dedicated research computers.

**Non-imaging data management.** REDCap study database on a HIPAA-compliant server, occur automatically according to a preset schedule (no participant action required). Data are immediately available for inspection and reporting of recruitment and retention status and any missing data. The study data management team performs weekly quality controls.

All datasets are cleaned, verified and archived. One official copy of all study data and a master data dictionary are maintained and updated regularly. All analytic and tracking databases are stored on a HIPAA-compliant server with continuous backups. For the protection of participant confidentiality, unique anonymous study IDs are used for data storing, tracking and reporting. PHI is stored separately from all other study data and will be used and disclosed in accordance with the HIPAA regulations. Regular reports are produced on (1) patient accrual and follow-up completion/retention in relation to goals and timeline; (2) the randomization process and group comparability on the balancing variables; (3) key baseline characteristics of the sample, by (blinded) group, related to the primary and secondary outcome variables; (4) intervention exposure and adherence; and (5) protocol violations. Any observed delays in these processes or data irregularities shall be followed up and resolved in a timely manner.

**Data sharing.** De-identified fMRI data and other study data are shared with collaborators at UIC and other collaborating institutions via a UIC Box Health Data Folder. All collaborating institutions are listed as data users under the HIPAA regulations and authorization provided by participants. PHI (e.g., name, address, phone number) and the link between study ID and the patients’ identities will not be shared. They can download data from the UIC Box Health Data Folder and store them on a HIPAA-compliant server at their institution for analysis according to the study protocol. We follow the latest industry standards for data encryption, server authentication, and client authentication to ensure secure data transmissions at all times. In addition, all the investigators and staff maintain up-to-date trainings and certifications in human subject’s protection, HIPAA, and Good Clinical Practice (GCP). We will strictly comply with the HIPAA and IRB regulation requirements regarding research use of PHI and appropriate safeguards for sharing coded data.

### 18.0 Data Analysis Plan and Sample Size

**Aim 1: Confirm neural target engagement**

***1.a. Primary Superiority Test***

In our primary superiority test, change in the Lumen Coached Group (G1) from week 0 to 18 is compared to change in the waitlist control group (G3) from week 0 to 18.^49^ The goal is to determine whether the Lumen group yields *greater improvement* than the waitlist control group. The measure will be constructed such that higher positive values are better; thus, change toward a *higher value is improvement*. The primary neural target is right dlPFC engaged by the go/nogo task. Secondary neural targets are left dlPFC engaged by the go/nogo task and bilateral amygdala engaged by negative facial emotion tasks.

The superiority test can be implemented as a between-group $t$-test on a change variable. We will construct a variable that includes the participants in both the Lumen and waitlist control groups that is defined as the final test (week 18) value minus the baseline value (week 0). When (mean) change in the Lumen group is greater than (mean) change in the waitlist control group, the Lumen group exhibits superior improvement compared to the waitlist control, or *superiority*. This will be evaluated in a 2-tailed statistical test as is recommended for superiority hypothesis testing in clinical research.^50^ The null hypothesis of no group difference in change may be rejected in favor of superiority when the observed $t$-test value for the difference in groups’ mean changes is *positive* and *exceeds the critical t value* for no group difference at $\alpha$=0.05 (2-sided).

***1.b. Secondary Noninferiority Test***

In the secondary noninferiority test, change in the Lumen Coached Group (G1) from week 0 to 18 is compared to change in the Human Coached Group^49,51^ (G2) from week 0 to 18. The goal is to determine whether the Lumen group yields *no less improvement* than the human-coached group. So the null hypothesis is that improvement in the Lumen group is below (less than) the improvement in the human-coached group by an amount as much as or greater than the *noninferiority margin* (Δ), whereas the alternative hypothesis is that the difference in improvement between the Lumen and human-coached group is smaller than the noninferiority margin.

H0: G2 – G1 ≥ Δ (G1 Lumen is inferior to G2 human coached PST by Δ or more)

H1: G2 – G1 < Δ (G1 Lumen is inferior to G2 human coached PST by less than Δ)

The noninferiority margin is a prespecified quantity to define how close the new treatment (Lumen) group mean must be to the active treatment control (human-coached PST) group mean to be considered not inferior. At present, there is no “gold standard” criterion for determining an appropriate noninferiority margin for the neural target measures in this study. Too large a margin would render rejecting the null hypothesis meaningless, whereas too small a margin would dramatically reduce the power to detect noninferiority.^52^ The confidence interval (CI) approach has often been used in both biomedical^53^ and mental health literatures,^54^ and regulatory authorities recommend the use of a 95% CI.^55,56^ Applying this approach, we choose the noninferiority margin to be a Cohen’s d=0.3 (small-to-medium effect), and if the upper 95% confidence limit for the standardized mean difference (G2-G1) is lower than d=0.3, then Lumen is considered not inferior to human-coached PST.

As described below, the primary superiority test is powered to detect a medium effect of d=0.45 for Lumen vs. waitlist control. Therefore, if the superiority test is positive then we expect the noninferiority test to be positively confirmed. This finding will indicate that Lumen is ready for confirmatory efficacy testing. However, it is possible that the superiority test is null but that the noninferiority test is positive. This finding will indicate that Lumen has promise and will be ready for confirmatory efficacy testing with further functional refinements to enhance its therapeutic efficacy. Furthermore, it is also possible that both the superiority and noninferiority tests are disappointing. This finding will indicate that Lumen will not be ready for confirmatory efficacy testing without further functional refinements, and likely also some redesign and additional pilot studies.

Once again the noninferiority test will be also based on a change variable and a between-groups $t$ test. In the superiority $t$-test for group mean differences in change, zero (0) represented the null hypothesis value which mean change has to reliably exceed in order to achieve superiority. In a noninferiority test the null hypothesis value that the mean change in the Lumen group must reliably exceed in order to achieve noninferiority is the human-coached group mean change minus (-) the noninferiority margin, expressed as G1 > G2 - Δ. If its mean change equals or falls below this value noninferiority cannot be claimed. As noted, the noninferiority test is in effect the same as a 1-sided superiority test for a special value (Δ). As such, the observed $t$ value can be calculated as mean change in the Lumen group plus (+) noninferiority margin, G1 + Δ, and compared with a critical $t$ value defined for no positive group difference. (The noninferiority margin is included in the observed $t$ rather than in the critical $t$ because the latter is always defined for a zero difference between groups.) Instead of testing that group difference in mean change > 0, we now test that group difference in mean change + noninferiority margin > 0. This allows the mean change in the Lumen group to be less than the mean change in the human-coached group, but not by more than the noninferiority margin.

Following published recommendation, we will conduct an ITT analysis with the goal of demonstrating noninferiority.^54,57^

**Aim 2: Examine the relationships of neural target engagement to outcomes**

In Aim 2, we will test 2 hypotheses: (1) both Lumen and human-coached PST will lead to greater improvements in PROs from week 0 to 18 compared with waitlist control; and (2) neural target engagement mediates these outcome changes. Additionally, we will compare changes in ecological daily assessments of mood, stress and coping over time between either PST group and waitlist control.

***2.a. Treatment effects on PROs***

Hypothesis 1 entails analyses that will compare differences in changes in PROs (symptoms, functioning, and quality of life) from week 0 to 18 across the 3 study groups. We will use Fisher's least significant difference (LSD) procedure, which is a two-step testing procedure: step 1: Analyses of ANOVA for the overall treatment effects on PROs, and step 2: if the step 1 is significant, then conduct further pairwise between-group *t*-test comparisons.^58^

***2.b. Mediating effects of neural targets on PROs***

The testing of mediation effects in the context of RCTs is now well established.^59-62^ Because change in the neural targets (mediator, M) is assessed at week 0 and 18, change in the PROs (outcome, Y) will be assessed at week 0 and 18 as well, so the focus is on contemporaneous mediation^63^ of neural target changes on changes in PROs before and after treatment. We will employ the approach recommended by Kraemer et al.,^64^ we defined that mediation exists if a potential mediator meets the two conditions. First, there is a significant effect of the intervention vs. control (X) on the potential mediator (M, X🡪M Path A). Second, the potential mediator is significantly associated with the outcome either as a simple effect in the usual care group or an interaction effect with the intervention relative to the usual care group (M🡪Y Path B). Our substantive hypothesis is that the mechanism of action of PST, delivered by Lumen or a human, is through activation of neural targets, which in turn alter depression and anxiety symptomatology and other PROs (e.g., psychosocial functioning and quality of life). If a future confirmatory efficacy trial is warranted based on data from this study, it can include multiple waves of follow-up assessments over a longer time to allow for testing temporal ordering of M and Y in longitudinal mediation analysis.

***2.c. Treatment effects on ecological daily assessments***

Longitudinal mixed modeling will be applied to analyses of the ecological daily measures (mood, stress, appraisal, and coping) where the design is Group$\times$Time=2$\times$8 as these measures will be obtained (using a validated measurement burst design)^65^ daily for 7 days every 2 weeks (on weeks 0, 2, 4, 6, 8, 10, 12, 18). During active treatment in the Lumen and human-coached groups, this will occur for 3 days prior to, the day of, and 3 days after each scheduled PST session. For waitlist controls, this will occur for 7 days starting on Sunday of each assigned week. This provides rich and detailed data on naturalistic changes over the course of treatment, as well as estimates of proximal responses to treatment within 3 days immediately post a PST session. For example, using the means of ecological measures over the 3 pre-PST days (to avoid confounding estimates with proximal treatment response) in the Lumen and human-coached groups and in the first 3 days in the waitlist control group every 2 weeks as dependent variables, we will characterize participant trajectories of change in terms of orthogonal linear, quadratic, and cubic components. We also will examine regression spline versions of these trajectories for visualization and interpretation. In a more exploratory vein, for each set of 7 daily ecological measures, collected biweekly, we will use piecewise linear models to characterize proximal responses to treatment based on the change of the 3 post-treatment days’ means from the 3 pre-treatment days’ means (omitting treatment day 4) in the Lumen and human-coached groups and the corresponding biweekly change of the last 3 days’ means from the first 3 days’ means (omitting day 4) in the waitlist control group. As these differences are nested within the 8 biweekly measurement bursts, and thus can be examined longitudinally, we can consider whether there is change over weeks in participants’ proximal responses to each PST session.

**Sample size calculation**

To determine the sample size, we power the study on the primary superiority test for the primary neural target in Aim 1, whereas our interest on superiority testing of the secondary neural targets and noninferiority testing of all neural targets in Aim 1 as well as all analyses in Aim 2 is to obtain effect estimates with reasonable precision and not to power in the traditional fashion for a desirable effect.

The primary contrast of interest (superiority test) involves only 2 of 3 treatment groups, comparing Lumen (G1) and waitlist control (G3). The contrast of secondary interest (noninferiority test) also involves only 2 groups, comparing Lumen (G1) and human-coached PST (G2). Note that these 2 contrasts are not independent, as they both involve the Lumen group, and this cannot be eliminated through any aspect of group sizing. For reasons of further development of Lumen, we decided to collect twice as much data for this experimental condition; thus, the allocation of participants is 2:1:1 for the Lumen, human-coached, and waitlist control groups, respectively.

For a research paradigm at the frontier of knowledge, a medium effect of 0.45 standard deviations by Cohen’s d standard is appropriate as this effect size screens out false-positive findings that could misdirect early-stage research. A sample of 150 (100 Lumen, 50 waitlist control) has 80% power to detect an effect of d=0.45 at α=5% (2-sided), assuming at least 85% retention.

As noted, the noninferiority test is secondary and our interest is to provide effect estimates with reasonable precision. In effect, the noninferiority test takes the same form as a 1-sided superiority test for the specified noninferiority margin (Δ=0.3).^66^ A sample of 150 (100 Lumen, 50 human-coached PST) at ≥85% retention provides 95% assurance that a precision interval with a 1-sided standardized half-width of 0.3 will contain the true effect parameter.

We also seek to provide reliable estimates of mediating effects of neural target engagement on outcomes in Aim 2. Fritz and MacKinnon (2007)^67^ demonstrated through simulations that the bias-corrected bootstrap standard error estimation of mediation effects—as provided in *Mplus* and some other programs—offers the greatest available power. In a suitable effect size metric, they examined all combination of treatment-to-mediator (X $\to$ M) α and mediator-to-outcome (M $\to$ Y) $\beta$ effects. In this metric, S=0.14 is *Small* (akin to Cohen’s d=0.20), H=0.26 is *Halfway* between Small and Medium (d=0.35), M=0.39 is *Medium* (d=0.50), and L=0.59 is *Large* (d=0.80). Thus, effect size combinations of HH, HM, MH, and MM mean $\alpha$ and $\beta$ parameters for the joint mediation path are in the small to medium range, which we consider reasonable given the exploratory nature of Aim 2. Sample sizes of 150 (100 Lumen, 50 waitlist control) and 100 (50 human-coached PST, 50 waitlist control) at ≥85% retention provide 95% assurance that precision intervals with 2-sided standardized half-widths of 0.35-0.45 will contain true effect parameters.

Therefore, we are confident of our ability to obtain reliable estimates of the effects between Lumen and human-coached PST for noninferiority testing as well as of the mediating effects. These effect estimates will be essential for informing future definitive trials that will be adequately powered to test noninferiority and mediation.

### 19.0 Provisions to Monitor the Data to Ensure the Safety of Subjects

This is a minimal risk study with prudent measures in place to protect the health and well-being of research participants. The following Data and Safety Monitoring Plan (DSMP) will be followed to ensure the safety of study participants and the validity and integrity of data in compliance with NIMH requirements.

**Independent Oversight.** Because the risks for adverse events (AEs) and data breaches are minimal in this RCT, and the study is not a Phase III clinical trial, it is determined that a Data and Safety Monitoring Board (DSMB) is not needed. Instead, a safety monitor *independent of* the study will be responsible for overseeing the implementation of this DSMP to ensure (1) the protection and safety of human subjects and (2) the validity and integrity of the trial. Dr. Bernice Man, MD, at the University of Illinois Chicago (UIC), serves in this role in the SPEAC-1 study and will continue to serve as a safety monitor in this study. Dr. Man is responsible for examining aggregate data on recruitment and retention, adverse events (unexpected or serious), and subject complaints, but she will not be reviewing individual, identifiable, subject data. If unexpected or serious adverse events occur, these will be reported to the IRB in the form of Prompt Reports. See further details below.

Contact PI Dr. Ma and research scientist Dr. Lv, both of whom will be blinded during the pilot RCT, will meet with Dr. Man every 6 months during active participant recruitment and follow-up to present and discuss data and safety monitoring information (below). Dr. Man will review and approve all meeting minutes. Annual continuing renewal applications to the IRB and annual progress reports to the NIMH will provide a summary of the discussions and recommendations from the meeting minutes regarding the following:

- ascertainment and any actions to be taken in response to AEs and SAEs reported during the study
- reports related to study operations and the quality of the data
- Possible modifications in the study protocol concerning recruitment, participant retention, data quality, or trial operations more generally.

**Safety Monitoring.** As in any clinical trial, it is not possible to anticipate all possible AEs. Staff will undergo extensive training in ascertaining, monitoring, and documenting AEs—serious or not. The study investigators have extensive experience in clinical trial organization and management, including data and safety monitoring for single site and multisite trials. Established procedures for rendering first aid and life-threatening emergencies will be monitored by Dr. Ajilore (psychiatrist).

An AE is defined as any untoward medical or psychological event experienced by a patient during or as a result of his/her participation in the study that represents a new symptom or an exacerbation of an existing condition whether or not considered study-related based on appropriate medical judgment. SAEs are any adverse experience that results in any of the following outcomes:

- Death
- Life-threatening event/illness
- Inpatient hospitalization or prolongation of existing hospitalization
- Persistent or significant disability/incapacity
- Pregnancy resulting in a congenital anomaly/birth defect
- Any event requiring medical or surgical intervention to prevent permanent impairment or damage. In this study, this is defined as physician confirmed diagnosis of any of the following: angina pectoris, heart attack, stroke, transient ischemic attack, heart failure, coronary angioplasty or bypass surgery, peripheral vascular disease, any other serious injury to the bone or muscle, liver failure, kidney failure, and cancer (except for non-melanoma skin cancer).

Non-serious AEs are all adverse events that do not meet the above criteria for “serious.” To ensure unbiased ascertainment, AEs will be systematically identified by querying participants at 18-week follow-up visits using the AE Patient Query Form. In order to access the EHR, as needed, for AE adjudication, consenting participants are asked to provide and/or confirm the minimum necessary identifiers including first and last name, date of birth, mailing address, and whether they have received care at UI Health.

Blinded Reporting. In this RCT, safety information will be monitored while keeping the true identity of the study groups masked. In his role as the study physician, Dr. Ajilore may need to become unblinded. But Contact PI Dr. Ma and biostatistician Dr. Xiao as well as the independent safety monitor Dr. Man will be blinded throughout the study and will review summaries of the numbers and rates of all AEs by blinded treatment group (treatment assignment of each group not revealed). Proper blinding of the other investigators and outcome assessors will be enforced as well.

Requirements for Adverse Event reporting. The Contact PI, Dr. Ma, will inform the Director of IRB panel and all relevant oversight committees at the university within 5 business days of learning of an unanticipated AE or major protocol deviation. All relevant information will be reported to the IRB for each unexpected SAE including information about the event and its outcome, dosing history of a suspect medication/treatment, concomitant medications, the participant’s medical history and current conditions, and all relevant laboratory data. Within 15 business days of the PI becoming aware of changes in risk/benefit or events requiring report to the sponsor, these will be reported. This timeline satisfies the NIMH reporting requirements for AEs and unanticipated problems. An annual report will be submitted to the IRB and to the sponsor summarizing all AEs, serious or not.

### 20.0 Provisions to Protect the Privacy Interests of Subjects

See Protection against breaches of participant privacy and confidentiality in Section 15.

### 21.0 Compensation for Research-Related Injury

There are no plans for the study to provide free medical care or to pay for research-related illnesses or injuries, or to provide other forms of compensation (such as lost wages or pain and suffering) to participants for research-related illnesses or injuries. As part of the informed consent process participants are advised to contact the PI to report any illness or injury experienced from taking part in the SPEAC-2 study. Additionally, participants who seek treatment through their regular doctor are advised to take a copy of the informed consent documentation which details all study assessment procedures and intervention activities.

### 22.0 Economic Burden to Subjects

There are no costs to participants for study devices or intervention. As part of the study participants receive online survey links and reminder notifications via text message on their mobile phone. The message and data rates will apply per their data plan.

### 23.0 Consent Process

**Recruitment.** Per HIPAA regulations, it is permissible to request a waiver of the Privacy Rule authorization requirement associated with the use or disclosure of PHI in patient medical records for the purpose of, and only for the purpose of, recruitment for clinical research studies. We have obtained IRB approvals of such requests in all of our past and current trials that use the same recruitment strategy by demonstrating our ability to satisfy the following criteria:

- The use or disclosure of the PHI involves no more than minimal risk to the privacy of individuals based on, at least, the presence of the following elements:
- An adequate plan to protect health information identifiers from improper use and disclosure.
- An adequate plan to destroy identifiers at the earliest opportunity consistent with conduct of the research (absent a health or research justification for retaining them or a legal requirement to do so).
- Adequate written assurances that the PHI will not be reused or disclosed to (shared with) any other person or entity, except as required by law, for authorized oversight of the research study, or for other research for which the use or disclosure of the PHI would be permitted under the Privacy Rule.
- The research could not practicably be conducted without the waiver or alteration.
- The research could not practicably be conducted without access to and use of the PHI.
- The PHI requested is the minimum necessary to achieve the purpose of the use or disclosure.

**2-Stage Informed Consent.**  All study participants involved in the SPEAC-2 trial must provide consent. A 2-stage process for initial screening consent and subsequent written informed consent will be followed to meet the ethical obligations to research participants and to foster a progressively increasing understanding of the research as well as the development of rapport between subjects and research staff. An online screening consent is first obtained prior to initial eligibility screening, which facilitates participant self-screening. Participant consent for eligibility screening is electronically documented in REDCap, as described in section 10, by selection of one of 3 options: 1) “I agree to participate, and I acknowledge that my selection of this option is intended to be the equivalent of my handwritten signature, or 2) I decline to participate or 3) I have read the above information and would like someone to call me with more information.” An electronic signature is not obtained for the screening consent. Second, screened eligible and interested participants are invited to attend a teleorientation with study staff who lead a full informed consent discussion and use the REDCap eConsent framework to provide written informed consent documentation. Study personnel obtaining informed consent are experienced in obtaining informed consent and receive standardized training in trial-specific protocols. Risks and benefits and the voluntary nature of the study are thoroughly explained by the study personnel. Participants have as much time and information as needed to consider whether or not to participate. The eConsent is obtained remotely via telephone or online video conferenced teleorientation session, as outlined in the enrollment procedures in Section 10.

**Informed Consent Process Training**. All study staff involved with the electronic informed consent process will receive uniform training and materials. These staff must demonstrate competency with simulated consent procedures prior to actual enrollment activities. A checklist will guide staff through electronic consent and data collection procedures to ensure compliance over time.

We will conduct effective and efficient informed consent to ensure that participants understand the following: (1) that the study is for research purposes and in no circumstances does it supplant medical care provided by their health care provider(s), (2) the risks and benefits of the study, (3) the available alternatives, and (4) that their voluntary decision to participate or to not participant in this research will be accepted without penalty (i.e. without jeopardizing their medical care or relationship with their health care provider). In reaching a decision about participation, it is essential for the potential subject to demonstrate an ability to use this information in a rational manner. Thus, in considering the risks, benefits, and available alternatives, subjects must show that they understand the aspects of these factors that are unique to them as individuals.

### 24.0 Process to Document Consent in Writing

All participants document their consent electronically using the secure REDCap eConsent framework and obtain their PDF copy of the signed consent by directly downloading through REDCap. Study staff confirms successful download with participant. As a backup, staff may use the REDCap **Send-It secure data transfer utility to allow participants to download the signed consent document securely by** emailing a unique download URL, along with a second email with the password to enable downloading the file. The study copy is automatically archived in the file repository in REDCap database, which is a HIPAA-compliant server, accessible to the PI and study personnel.

### 25.0 Setting

Participants will be recruited from UIC and UI Health and the local areas. The assessment visits, including fMRI are conducted at the 3T MR Research Program located at 2242 W Harrison, the UIMC Advanced Imaging Center, a BioRAFT registered facility. The Lumen Orientation Visit and the first PST session in the human-coached intervention will take place at UIC’s Westside Research Building located at 1747 W Roosevelt Road.

**Appendix 1.	SPEAC-2 Schedule of Measures**

| **Measure** | **Instrument** | **Collection Method** |  | **Time** (weeks)**^a^** | | |
| --- | --- | --- | --- | --- | --- | --- |
|  |  |  | **Screening** | **0** | **18** | **1-12** |
| Eligibility criteria | Age, sex, current/planned pregnancy/lactation, etc. | Self-report | **X** |  |  |  |
| Depression severity | Patient Health Questionnaire-9 (PHQ-9), suicidal ideation (item #9) | Self-report | **X** |  |  |  |
| Anxiety | Generalized Anxiety Disorder Scale (GAD-7) | Self-report | **X** |  |  |  |
| Alcohol/substance abuse | CAGE Adapted to Include Drugs (CAGE-AID) | Self-report | **X** |  |  |  |
| Brain scan screening | Brain scan screening questions (e.g., self-reported weight) | Self-report | **X** |  |  |  |
| Cognitive Impairment | Callahan 6-item screener | Interview | **X** |  |  |  |
| Emotional reactivity and cognitive control | fMRI | Measured |  | **X** | **X** | NA |
| Emotional reactivity and cognitive control | Penn State Worry Questionnaire^68^  Positive and Negative Affect Schedule (PANAS)^69^ | self-report |  | **X** | **X** | NA |
|  | Social Problem Solving Inventory-Revised: Short (SPSI-R:S)^70^  Dysfunctional Attitudes Scale^71^ | self-report |  | **X** | **X** | NA |
| Depression and anxiety symptoms | Hospital Anxiety and Depression Scale (HADS)^72,73^ | self-report |  | **X^b^** | **X^b^** | NA |
| Functioning | Sheehan Disability Scale,^74^ Work productivity and activity impairment questionnaire (WPAI)^75^ | self-report |  | **X** | **X** | NA |
| Quality of life | 12-item Short-Form Health Survey (SF12)^76^ | self-report |  | **X** | **X** | NA |
| Mood | Daily mood (items from circumplex model, capturing affective valence and arousal) | Ecological end-of-day assessments, per Event Window table in Appendix 4. |  | **X** | **X** | **X^c^** |
| Stress | Daily stress events (15 categories), perceived stressor severity, stressor-related thoughts |  |  | **X** | **X** | **X^c^** |
| Appraisal | Assessing daily implementation of “problem orientation” (challenge appraisals, optimism, self-efficacy, outcome expectancies) |  |  | **X** | **X** | **X^c^** |
| Coping | Assessing daily implementation of “problem solving-style” (attempts to understand problems, plan effective solutions to coping, impulsivity, carelessness, avoidance behaviors) |  |  | **X** | **X** | **X^c^** |
| Depression and anxiety symptoms | PHQ-9 and GAD-7 | Self-report and data tracking during PST sessions |  | **X^b^** | NA | **X^b^** |
| Treatment acceptability | Session metrics: Number and duration of PST sessions completed; End-of-session Lumen user survey (after S1, S4, S8): User Experience Questionnaire-Short version (UEQ-S),^77^ adapted Working Alliance Inventory for digital interventions (WAI-Tech)^78,79^ |  |  | NA | NA | **X** |
| Digital health literacy | Digital Health Literacy Instrument^45^ | self-report |  | **X** | NA | NA |
| Experiences and discrimination | PhenX Major Experiences and Everyday Discrimination Scales^80^ | self-report |  | **X** | NA | NA |
| Height | Height | Measured |  | **X** | NA | NA |
| Weight | Body weight | Measured |  | **X** | **X** | NA |
| Blood pressure | Blood pressure | Measured |  | **X** | **X** | NA |
| Sociodemographics | age, sex, race, ethnicity, education, race, ethnicity, income, household size, marital status, employment status, occupation, smoking/vaping | self-report |  | **X** | NA | NA |
| Adverse Events (AE) | AE form | Interview |  | **X** | **X** | NA |

**^a^**To minimize missing data, participants will receive $50 at wk 0, study iPad (unlocked) or $100 (if opting out iPad) at wk 18, and $1 per daily diary.

**^b^**HADS is used as the independent outcome measure of depressive and anxiety symptoms at weeks 0 and 18. PHQ-9 and GAD-7 are used for 2 purposes: (1) eligibility screening (before week 0) and (2) treatment progress monitoring across PST sessions (in weeks 1, 2, 3, 4, 6, 8, 10, 12).

**^c^**In addition to ecological daily assessments that will occur for 7 days at weeks 0 and 18, they also will occur every 2 weeks (on weeks 2, 4, 6, 8, 10, 12). During active treatment in the Lumen coached arm (Studies 1 & 2) or the human coached PST arm (Study 2), this will occur for 3 days prior to, the day of, and 3 days after each scheduled PST session. For waitlist controls, this will occur for 7 days starting on Sunday of each assigned week.

**Appendix 2. Brief Description of Study Outcome Measures**

| **Type** | **Name** | **Brief Description** |
| --- | --- | --- |
| Primary | Functional magnetic resonance imaging (fMRI) | Outcome Description Two neural targets defined a priori, specifically activation of the amygdala for nonconscious threat-related emotional reactivity and activation of the DLPFC for cognitive control will be assessed using fMRI. In the facial emotion viewing paradigm, facial expression stimuli are standardized black and white photographs of 8 identities (4 female, 4 male) with evoked expressions of threat-related emotions (fear, anger), loss-related emotions (sadness) and reward-related emotions (happiness), along with neutral. For the Go-NoGo paradigm, the 'Go' and 'NoGo' stimuli are presented for 500 ms each with an inter-stimulus interval of 750 ms. The 2 paradigms are empirically validated probes of emotional reactivity and cognitive control, respectively. |
| Secondary | Penn State Worry Questionnaire (PSWQ) | Outcome Description PSWQ is a self-reported, 16-item, Likert-type scale that measures the trait of worry. Cronbach's alpha is 0.93-0.95 and test-retest correlation is 0.92 after 8-10 weeks. In addition, PSWQ significantly discriminates levels of generalized anxiety disorder (GAD) and GAD versus PTSD. |
| Secondary | Positive and Negative Affect Schedule (PANAS) | Outcome Description PANAS consists of two 10-item self-reported scales to measure positive and negative affect. Each item asks about the extent one has felt a positive or negative feeling on a 5-point scale of 1 (not at all) to 5 (very much). Cronbach's alpha is 0.86-0.90 for the positive affect items and 0.84-0.87 for the negative affect items when reporting affect across time frames from at this moment to a year. Test-retest correlation is 0.68 for positive affect and 0.71 for negative affect when reporting affect in general over an 8-week period. |
| Secondary | Social Problem-Solving Inventory-Revised: Short Form (SPSI-R:S) | Outcome Description Participants' problem solving abilities will be assessed using the reliable and valid SPSI-R:S that contains 25 items in the following 5 subscales: positive problem orientation (PPO), negative problem orientation (NPO), rational problem solving (RPS), impulsive/careless style (ICS), and avoidance style (AS). Each item is rated on a 5-point scale ranging from "not at all true of me" (0) to "extremely true of me" (4). SPSI-R:S is a short version of the long SPSI-R:L form which has Cronbach's alpha coefficient of 0.95 for the total score and of 0.67-0.92 for subscales. |
| Secondary | Dysfunctional Attitudes Scale (DAS) | Outcome Description DAS (Form A) is a 40-item self-reported scale that measures the presence and intensity of dysfunctional attitudes. Each item is rated a 7-point Likert scale (7 = fully agree; 1 = fully disagree). The higher the sum of the 40-items, the more dysfunctional attitudes an individual possesses. Cronbach's alpha is 0.89-0.92 and test-retest correlation is 0.73 over a 6-week period. DAS was found to be significantly correlated with the Beck Depression Inventory in an adult population (r=0.41). |
| Secondary | Hospital Anxiety and Depression Scale (HADS) | Outcome Description HADS, including 7 questions for anxiety and 7 questions for depression, measures self-reported anxiety and depression in a general medical population of patients. Each item on the questionnaire is scored from 0-3 and the score for either anxiety or depression ranges between 0 and 21 with the following categories: normal 0-7, mild 8-10, moderate 11-14, and severe 15-21. Total score for the entire scale (emotional distress) ranges from 0 to 42, with higher score indicating more distress. Cronbach's alpha coefficients range from 0.78-0.93 for the anxiety subscale and from 0.82-0.90 for the depression subscale. Test-retest correlations are ≥0.80 after ≤2 weeks. Correlations of the anxiety and depression subscales with commonly used anxiety and depression measures (e.g., Beck Depression Inventory, Patient Health Questionnaire, State-Trait Anxiety Inventory, Symptom Checklist-90-Revised) vary between 0.60 (good) and 0.80 (very good). |
| Secondary | Sheehan Disability Scale | Outcome Description The Sheehan Disability Scale is a validated questionnaire that measures functional disability and is sensitive to treatment effects in clinical trials. Cronbach's alpha is 0.89. Patients rate the extent to which their symptoms impair work/school, social, and family life on a visual analog scale from 0 to 10 and answer the number of days when their symptoms cause them to miss work/school and be unproductive at work/school. |
| Secondary | Work productivity and activity impairment questionnaire (WPAI) | Outcome Description The WPAI was created as a patient-reported quantitative assessment of the amount of absenteeism, presenteeism and daily activity impairment attributable to general health in the past 7 days. All measures of work productivity and activity impairment were positively correlated with measures which had proven construct validity. These validation measures explained 54 to 64% of variance (p less than 0.0001) in productivity and activity impairment variables of the WPAI. |
| Secondary | 12-item Short-Form Health Survey (SF12) | Outcome Description The SF-12 is a 12-item version of the SF-36 that measures overall health-related quality of life. Physical and mental health composite scores are computed using the scores of 12 questions and range from 0 to 100, with 0 indicating the lowest level of health and 100 indicating the highest level of health. Test-retest correlation is 0.89 for the physical health subscale and 0.76 for the mental health subscale. |
| Secondary | Daily Mood | Outcome Description Daily end-of-day assessments will be captured over a one-week period at baseline and every 2 weeks (i.e., on weeks 2, 4, 6, 8, 10, 12, and 18). During active treatment in the Lumen arm (Studies 1 & 2) or the in-person PST arm (Study 2), this will occur for 3 days prior to, the day of, and for 3 days after a scheduled PST session. For waitlist controls, this will occur for 7 days starting on Sunday of each assigned week. Daily mood will be measured using 8 mood items that capture both the arousal and valence components of the circumplex model of affect. |
| Secondary | Daily Stress | Outcome Description Daily end-of-day assessments will be captured over a one-week period at baseline and every 2 weeks (i.e., on weeks 2, 4, 6, 8, 10, 12, and 18). During active treatment in the Lumen arm (Studies 1 & 2) or the in-person PST arm (Study 2), this will occur for 3 days prior to, the day of, and for 3 days after a scheduled PST session. For waitlist controls, this will occur for 7 days starting on Sunday of each assigned week. Daily stress will be assessed by report of stressors on that day; when stress is reported, the nature/characteristics of the stressor and response will be assessed. The type of stress will be assessed using a list of 12 categories of social problems, followed by reporting the perceived severity of the most salient stressor that day, and the degree to which the participant has had stressor-related thoughts (e.g., intrusive thoughts). Finally, we will capture an overall perceived stress report for each day (regardless of the number and type of stressors reported). |
| Secondary | Daily Appraisal and Coping | Outcome Description Daily end-of-day assessments will be captured over a one-week period at baseline and every 2 weeks (i.e., on weeks 2, 4, 6, 8, 10, 12, and 18). During active treatment in the Lumen arm (Studies 1 & 2) or the in-person PST arm (Study 2), this will occur for 3 days prior to, the day of, and for 3 days after a scheduled PST session. For waitlist controls, this will occur for 7 days starting on Sunday of each assigned week. Two components of these assessments will capture a summary of appraisal and coping processes associated with daily experiences. The appraisal items will focus on assessing "problem orientation" or the general awareness and appraisals of problems during the day. The coping items will focus on assessing "problem-solving style" or the methods of planning and coping with the most salient problem experienced during the day. |
| Other | Patient Health Questionnaire (PHQ-9) | Outcome Description PHQ-9 is a self-administered instrument for screening, diagnosing, monitoring, and measuring the severity of depression. It rates the frequency of symptoms as "0" (not at all) to "3" (nearly every day) and has been validated for use in primary care. The PHQ-9 total score ranges from 0 to 27 and is categorized as follows: None or minimal depression 0-4, Mild depression 5-9, Moderate depression 10-14, Moderately severe depression 15-19, and Severe depression 20-27. Cronbach's alpha coefficients range from 0.86 to 0.89 and test-retest correlations range from 0.84-0.95 within 48 hours and from 0.81-0.96 at 7-day reassessment. PHQ-9 scores were found to be highly correlated with Beck Depression Inventory scores in the general population (r=0.73). |
| Other | Generalized Anxiety Disorder Scale (GAD-7) | Outcome Description GAD-7 is a valid and reliable 7-question scale to screen for 4 anxiety disorders: Post Traumatic Stress Disorder, Panic Disorder, Generalized Anxiety Disorder, and Social Phobia. A score of ≥10 indicates a high probability of 1 or more of these disorders. Cronbach's alpha is 0.92 and test-retest correlation is 0.83. GAD-7 scores also correlate highly with scores of 2 commonly-used anxiety scales: the Beck Anxiety Inventory (r=0.72) and the anxiety subscale of the Symptom Checklist-90 (r=0.74). |
| Other | User Experience Questionnaire-Short version (UEQ-S) | Outcome Description UEQ-S is a validated instrument containing 8 items from the 26 items of the original UEQ. The 8 items are grouped into 2 subscales, pragmatic and hedonic quality (4 items each). A total value reflects the overall user experience. Cronbach' alpha values are 0.85 for the pragmatic quality subscale and 0.81 for the hedonic quality subscale. |
| Other | Adapted Working Alliance Inventory for digital coaching interventions (WAI-Tech) | Outcome Description WAI-Tech was based closely on the original 36-item WAI and uses a parallel set of 36 items rated on a 7-point scale (1 = "never" to 7 = "always") to measure the level of alliance between patient and digital coach along 3 domains: Task (12 items), Bond (12 items), and Goal (12 items). The Task subscale measures how responsive the digital coach was to the patient's focus or need. The Bond subscale measures the degree of the affective bond between patient and digital coach. The Goal subscale measures the extent to which goals were important, mutual, and capable of being accomplished. The WAI-Tech yields 3 subscale scores and 1 overall score. Cronbach's alpha coefficients are 0.92 for the total scale, 0.84 for the Task subscale, 0.78 for the Bond subscale, and 0.75 for the Goal subscale. |

**Appendix 3.** **Study Event Windows**

**Event windows** (in Days)

| **Preferred windows (in Days)** | **IES** | **V1** | **EMA**#**1** | **R** |  |  |  |  |  |  |  |  |  |  |  |  |  |  | **V2** |
| --- | --- | --- | --- | --- | --- | --- | --- | --- | --- | --- | --- | --- | --- | --- | --- | --- | --- | --- | --- |
| Lumen or human-coached PST: |  |  |  |  | **Lumen Orient.** | **S1** | **S2** | **S3** | **S4** |  | **S5** |  | **S6** |  | **S7** |  | **S8** |  |  |
| **Week (indexed from randomization) #:** from  to | -4  -2 | -2  -1 |  | 0 | 1  2 | 1  3 | 2  4 | 3  5 | 4  6 |  | 6  8 |  | 8  10 |  | 10  12 |  | 12  14 |  | **16**  **20** |
| IES-eligible to eConsent, max. days | 14 |  |  | **Randomization** |  |  |  |  |  |  |  |  |  |  |  |  |  |  |  |
| eConsent to V1, max. days |  | 14 |  |  |  |  |  |  |  |  |  |  |  |  |  |  |  |  |  |
| *IES-eligible to V1* *(rescreen if IES-V1>90d), max. days* | 28 | |  |  |  |  |  |  |  |  |  |  |  |  |  |  |  |  |  |
| V1 to EMA#1 start (automated), max. days |  | **1** |  |  |  |  |  |  |  |  |  |  |  |  |  |  |  |  |  |
| V1 to Randomization, max. days |  | **10** | |  |  |  |  |  |  |  |  |  |  |  |  |  |  |  |  |
| Randomization to Lumen Orientation Visit, max. days |  |  |  |  | 14 |  |  |  |  |  |  |  |  |  |  |  |  |  |  |
| Lumen Orientation to Session 1, max. days |  |  |  |  |  | 7 |  |  |  |  |  |  |  |  |  |  |  |  |  |
| Randomization to human-coached Session 1, max. days |  |  |  |  | N/A | 14 |  |  |  |  |  |  |  |  |  |  |  |  |  |
| Lumen and human-coached Session to Session, max. days |  |  |  |  |  |  | 7 | 7 | 7 | 14 | | 14 | | 14 | | 14 | |  |  |
| Randomization to V2, target 126d (18 weeks) +/- allowable 14d window (16-20 weeks) |  |  |  |  |  |  |  |  |  |  |  |  |  |  |  |  |  |  | 126  +/-14 |

### Abbreviations: IES, initial eligibility screening; EMA, ecological daily assessment; PST, problem solving treatment; S1-S8, sessions 1-8 with Lumen; V1, visit 1; V2, visit

### Section B: Inclusion and Exclusion Criteria

*Inclusion Criteria*

- Age: ≥ 18 years
- Emotional distress defined by elevated depressive (PHQ-9 scores 10-19) and/or anxious symptoms (GAD-7 scores 10-14)
- Willing and able to provide informed eConsent and HIPAA authorization

*Exclusion Criteria*

- Unable to speak, read, or understand English for informed consent
- Current pharmacotherapy or psychotherapy (individual or professionally led group therapy) for depression or anxiety (note: participants are not withdrawn post-randomization if they begin pharmacotherapy drugs or start psychotherapy during the study.)
- Suicidal ideation per PHQ-9 with active plan
- Bipolar or psychotic disorder, or current psychiatric treatment
- Weight ≥325 pounds due to brain scanner constraints, MRI contraindications, traumatic brain injuries, and tumor or any other known structural abnormality in the brain
- Severe medical condition (e.g., myocardial infarction or stroke or new cancer diagnosis in the past 6 months, end-stage organ failure, terminal illness) or residence in a long-term care facility
- Diagnosis of cancer (other than non-melanoma skin cancer) that is/was active or treated with radiation or chemotherapy within the past year
- Active alcohol or substance use disorder (including prescription drugs) based on the CAGE Questionnaire Adapted to Include Drugs (CAGE-AID)
- Cognitive impairment based on the Callahan 6-item screener
- Current or planned pregnancy or lactating (<6 months postpartum)
- Participation in other investigational treatment studies that would significantly affect participation in this study, raise safety concerns, and/or confound outcomes (participant may be asked to provide the informed consent of the other study for final decision on exclusion by a study psychiatrist)
- Family/household member of an already enrolled participant or of a study team member
- Plan to move out of the Chicago area during the study period
- Does not have reliable Wi-Fi Internet at home
- Unwillingness to use personal mobile device to receive study text messages
- Investigator discretion for clinical safety or protocol adherence reasons

### Section C: Lumen Architecture

Lumen is a virtual voice-based coach that delivers Problem Solving Treatment (PST) to counsel participants with depression and/or anxiety using the 7-step problem solving process and the SSTA (stop, slow down, think and act) method of coping. Lumen conducts interactive conversations providing appropriate responses based on participant input. In order to deliver seamless interactions, we developed a robust and parsimonious architecture that combines PST, integrated components to provide context for Lumen conversations, data storage for the interactions, and a software infrastructure for providing security and privacy for these interactions.

Lumen architecture (see **Figure S1**) was developed with input and consultation from researchers (in computer science, medicine, health service researchers, and psychiatry), software developers, and human computer interaction experts. The architecture was developed on Amazon’s Alexa platform, with further integration with a secure REDCap system for ecological momentary assessments (EMAs) and surveys. Lumen’s software architecture is comprised of two components: a conversation manager module and a context manager module.

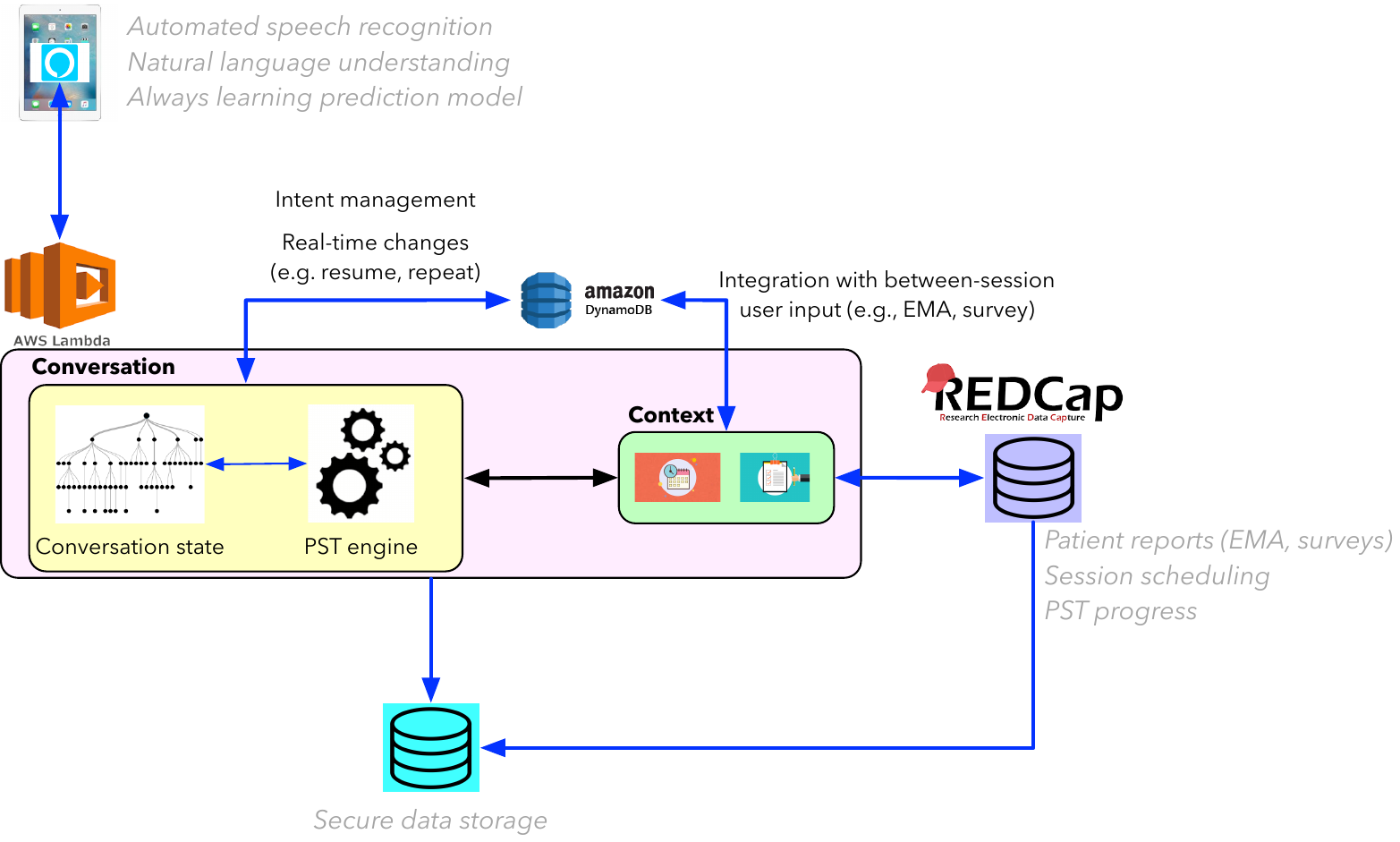

**Figure S1. Lumen software architecture that includes the conversation manager and context manager modules.**

The **conversation manager** is the voice-interactive component of Lumen and is responsible for delivering PST content. This PST content is aligned with the theoretical constructs and associated treatment guidelines. Towards this end, the conversation manager includes a PST engine that incorporates PST-related and conversational structures. The PST engine is a flexible set up that allows for the adaptation of PST content for other health-related problems (e.g., translating the therapy for weight management or smoking cessation) in the future.

Within the Lumen application, the PST engine tracks the progress of a PST session. For example, in a session, once the participant defines a specific problem to work on (step 1), Lumen will proceed to guide the participant in establishing a realistic goal for solving the problem (step 2). Then, once the participant has defined a set of potential solutions that can meet the goal (step 3), Lumen will guide the participant on evaluation of the pros and cons of each solution (step 4). After the participant chooses his/her preferred solution (step 5), Lumen will guide the participant to develop an action plan (step 6), which Lumen will prompt the participant to implement and evaluate the outcome post the session (step 7). This stepwise structure is the same in all PST sessions as is the case in current practice, and the participant identifies a problem to work on in each session (except session 1, which is an introductory overview session situating the PST process and building an initial problems list which participants can choose to work on in subsequent sessions, or they can opt to work on a new problem). Importantly, each of these steps is based on established PST theory and practice.

In addition, the conversation manager also manages the “state” of the conversation—including the flow of interactive conversation (e.g., the flow of the above-mentioned steps), and adaptive responses based on user input. The conversational state is adaptively managed based on user input. Towards this end, user “intents” or statements are parsed and mapped into pre-defined categories, which are then mapped to the appropriate state in the PST interaction to deliver relevant content, aligned with the user input. The conversation manager also provides dynamic support for functions such as resume, which allows participants to re-start a previously incomplete PST session. Such stop-restart functions provide flexibility in conversational interactions, affording perceptions of realistic conversational interactions.

The conversation manager also interacts with a **context manager** module that provides situated and contextual information regarding Lumen sessions. The context manager primarily controls three aspects: persistence of therapy content across sessions, scheduling/re-scheduling of sessions, and integrating external content (e.g., surveys, EMAs) into the Lumen sessions. The context manager dynamically tracks user identified problems (see step 1 above), and goals that were previously developed and asks participants to self-evaluate adherence to their previously developed action plans in ensuing sessions. Such dynamic follow-up increases the persistence and continuity of therapy across sessions, developing trust and confidence in the virtual coach. Similarly, the context manager tracks session progress (e.g., session 5), and helps participants schedule and re-schedule sessions. With its integration with an external scheduling database (in REDCap), follow-up emails and messages are tracked to ensure that the therapy sessions are synchronized with participant needs. Finally, the context manager also interacts with the REDCap database to incorporate survey responses (e.g., completion of the PHQ-9 and GAD-7 surveys before each session) into the Lumen session. For example, prior to the start of each Lumen session, participants are asked to complete the PHQ-9 and GAD-7 surveys; if incomplete (or partially complete), Lumen prompts the user to complete the surveys then re-start the session.

All sessions and communicative interactions are stored in a secure AWS-based database for analysis. In order to prevent accidental recording, and for pragmatic implementation in a clinical trial, we currently have implemented the entire Lumen infrastructure in a “locked down” mode in an 8^th^ generation Apple iPad. Participants can access the Lumen application skill within the Alexa application, preloaded on their study iPad, by stating “Alexa, Open Lumen….” A summary of the Lumen components is provided in Table S1.

**Table S6. Lumen components, and their associated functions**

| **Lumen Component** | **Functions** |
| --- | --- |
| Conversational manager | Managing conversations based on user intent, aligning with PST constructs, additional functions to manage conversations (e.g., repeat, resume), tracking progress within a session |
| Context manager | Tracking user problems and goals (from previous PST sessions), prompting participants to self-evaluate action plan adherence, integrating user responses from PHQ-9 and GAD-7 surveys within sessions, scheduling/re-scheduling sessions, |
| Lumen REDCap database | Ecological daily assessments, patient surveys, patient calendaring and reminders |
| Storage/Data Management (AWS & REDCap) | Comprehensive storage of user responses to surveys, user interactions with the Lumen, synchronization across devices, data security and privacy |

### Section D: Participant Interactions with Lumen

Lumen architecture that is presented in Section B, is realized through user interactions with the Lumen skill embedded within the Alexa application (currently delivered on an iPad device) and user completion of surveys and EMAs with hyperlinks delivered via emails and text messages (on the participant mobile phones).

Participant interaction with the Lumen PST coach involves two primary components: (a) sessions with the coach, and (b) completion of surveys, EMAs, administrative management (e.g., session scheduling, re-scheduling, monitoring progress on their personal dashboard).

Participants access their scheduled Lumen session via the Alexa application on their assigned iPad. Sessions are instantiated by the participant by saying “Alexa, Open Lumen.” Conversations then continue based on the progress that participants have made (e.g., number of sessions completed), current problem(s) being addressed and implementation of previously created goals and action plans. As previously described, these conversations are aligned with PST’s treatment protocol and guidelines and with conversational structures and flow that reinforce the patient-centered approach of PST. As with regular conversations, participants can exit, resume or ask the Lumen coach to repeat parts of the conversations that they are unable to follow. At the end of each session, participants schedule their next session with Lumen.

Lumen’s PST sessions are delivered on a dedicated iPad, which is encrypted and functions in a “lockdown” mode with no additional functions or local storage. The purpose of using such an approach is for trialing it in a controlled environment without compromising on data safety and privacy of the user. Additionally, such an infrastructure and set-up provide several advantages. First, it creates the perception of a “device as a coach” mode, assigning a specific role-based purpose (i.e., Lumen as a health coach, that is delivered only on the specific device). Second, embedding the Lumen skill within the Alexa application reduces the potential for accidental recording (as is common with smart speaker devices such as Echo or Google Assistant). Finally, the encrypted mode with no local data storage allows for data privacy protection and remote management, in case of a lost or misplaced device.

Between sessions, participants will also complete surveys and EMAs to track their progress. These administrative tasks and surveys, which provide situated context for Lumen interaction and monitoring of progress will be delivered on the participant’s personal devices. The surveys and EMAs are sent by text message with embedded links. Participants complete these on any browser associated with their mobile phone. Additional messages will include reminders about sessions, ability to schedule/re-schedule planned sessions, and other session related information.

All of the survey, EMA and scheduling tasks are instantiated through a dedicated REDCap project. Based on an initial session date, a program calendar according to PST guidelines (4 weekly, then 4 biweekly sessions) is set up and used for all session reminders. Future session appointments are confirmed or adjusted per user preference during Lumen sessions and are automatically updated in REDCap to assure that PHQ-9 and GAD-7 surveys are delivered at appropriate times.

### Section E: Components of PST Implemented in Lumen

To align with the treatment fidelity of the evidence-based PST, components of PST were implemented within the conversation manager and context manager of Lumen. The timing of the delivery of these components were aligned with the evidence-based PST. Each of the PST components, the timing of the delivery (i.e., session), and the aspects of the component managed by the conversation manager and context manager are provided in Table S2.

**Table S7. Components of PST implemented within the conversation and context managers of Lumen**

| **PST Fidelity Components** | **Session** | **Lumen Conversation Manager** | **Lumen Context Management (survey, EMA, participant workbook)** |
| --- | --- | --- | --- |
| **Introduction to Lumen and Building Rapport** | **Session 1** | Lumen introduces and tells participants about its role and purpose Personalized information | • Utilizes participant specific information stored in a profile |
| **Introduction to the Program and PST Overview** | **Session 1** | Natural conversation and verbal description of Lumen, background on PST and how it works |  |
| **Administer and Verify Completion of PHQ-9 and GAD-7** | **All Sessions** | Lumen checks and validates if surveys are completed within past 3 days, and, if not completed, provides directions on how to complete them | • REDCap links to complete PHQ-9/GAD-7 surveys externally sent via SMS. • Survey reminders sent before each session  • If session with Lumen begins and the PHQ-9/GAD-7 surveys are not complete, participant is asked to exit the session, complete surveys, then resume session • If PHQ-9/GAD-7 surveys are not completed within 3 days of completing Lumen session, participants sent new link and asked to re-take surveys. • REDCap links to complete brief (2-3 minute) EMAs that assess mood are sent for 7 days in a row, every other week. EMAs are open from 7PM to midnight on the day they are sent. If an EMA is not filled out by 9:30PM on the day it is sent, participants will receive one reminder to complete it. |
| **Knowledge Check** | **Session 1** | Multiple Choice questions asked by Lumen to evaluate participant understanding of key concepts before problem solving begins (e.g., link between unresolved problems and emotional distress, number of skills to be learned in the program, following a person-centered approach, rationale for utilizing PHQ-9/GAD-7, behavioral activation and planning varied activities) |  |
| **Problem List Generation** | **Session 1** | Lumen prompts participants to list problems they would like to work on in subsequent sessions | • Participants fill out the "Problem List" page in their workbook. |
| **7-Step PST Process** | **Sessions 2-8** | Lumen verbally walks participants through the steps of problem solving (Problem Identification, Goal Selection, Generating Possible Solutions, Solution Evaluation, Solution Choice, Action Plan Development). Lumen confirms (by repeating) participant answers and guides them to self-evaluate their responses at each step. Participants are asked if they need additional time as they complete each step.  Lumen will recall selected problems and goals and will ask participant about their action plan implementation at the next session as part of the progress review. | • Participants fill out "Problem Solving Worksheet" in their workbook to keep track of their listed problems. |
| **Progress Review** | **Sessions 3-8** | Prior to starting a new problem-solving session, Lumen asks participants to evaluate their action plan implementation from the immediate previous session  Lumen reminds participants of problems and goals they’ve chosen in the previous session. Further follow up is also conducted regarding how the participant was able to implement and adhere to the action plan, with Lumen prompting the participant to self-evaluate their action plan adherence. Additional questions regarding whether planned activities (Behavioral Activation) were carried out are also asked. | • Fill out "Progress Review" section of the workbook |
| **Behavioral Activation** | **Sessions 2-8** | Lumen explains 3 activities (social, physical, pleasant), their importance, along with examples. Participants are guided through planning 1 of each type of activities to partake in by the next session.  Lumen confirms participant activity choice and will ask participants whether they were able to complete their scheduled activities, at the next session, as part of the progress review | • Fill out "Behavioral Activation Worksheet" section of the workbook |
| **Scheduling, Rescheduling, Reminders** | **Between each session** | Participants are sent text message reminders about upcoming Lumen sessions (2/day) and asked to complete PHQ-9 and GAD-7 surveys before starting their upcoming Lumen session.  The timing of the sessions are based on session 1 (Lumen asks participant to write these down on the "Lumen Sessions" page in their workbook). Participants are encouraged to follow the pre-arranged session schedule; in case of emergency, participants have the ability to reschedule their Lumen session to a day before or after their pre-arranged session schedule. |  |

**Note that the contents in this supplementary material Sections A through C, have been previously described in the primary Lumen design paper ^40^ and the pilot trial paper.^81^

### Section F: Neuroimaging Acquisition and Processing Protocol

Imaging Sequences

BOLD contrast functional images were acquired with echo-planar T2*-weighted imaging using a GE MR750 3T scanner (GE Healthcare, Milwaukee, Wisconsin) with a NOVA 32-channel head coil. Head motion was restricted with foam pads.

Each whole brain volume consisted of 45 interleaved 3mm thick axial/oblique slices (74 x 74 matrix; TR=2000ms; TE=27.5ms; voxel size=3x3x3mm; FOV=222mm; flip angle=77°). One hundred fifty-four volumes were acquired over 5 minutes and 8 seconds for both tasks. A high-resolution T1-weighted structural scan was acquired using GE’s BRAVO sequence at the end of the imaging session for use in normalization of the fMRI data into standard space with the following parameters: TR=0.008, TE=0.003; voxel size=1x1x1mm; number of slices=176; FOV=256x256; flip angle=11^o^.

Image Pre-processing

Pre-processing and data analysis were performed using Statistical Parametric Mapping (SPM) software implemented in Matlab (SPM8; Wellcome Department of Cognitive Neurology) and the FSL^82^ in a manner similar to that of our prior publications.^83,84^ Briefly, motion correction was performed by realigning and unwarping the fMRI images to the first image of each task run after removal of the three dummy scans acquired at the start of the scanning session. Images were normalized to the stereotactic space of the Montreal Neurological Institute template.^85^ T1-weighted data were normalized to standard space using the FMRIB nonlinear registration tool, and the functional echo-planar image data were co-registered to the T1 data using the FMRIB linear registration tool. Prior to computing brain activation values, physiological noise was estimated using the time series from an eroded mask within the ventricles and white matter and was removed from the motion-corrected fMRI time series. Functional data were then smoothed using an 8 mm Gaussian kernel and high-pass filtered using a cutoff period of 128 seconds.

Following realignment and unwarpinboxg, quality control diagnostics were completed on the time series data for each run. Quality control diagnostics included removing scans with incidental findings, scanner artefacts and signal dropout. Participants’ data were included if no more than 25% of time points were censored for frame-wise displacement or variance spikes.

Defining regions of interest

Our target regions of interest are activation of the dlPFC for cognitive control (primary) and activation of the amygdala for nonconscious threat-related emotional reactivity (secondary). These regions were defined a priori.^86,87^ Pre-planned analytic plan was established in a prior systematic procedure validated with the same tasks as used in the present trial.^86^ Our a priori focus on these regions was informed by our prior studies. The SPEAC-1 pilot RCT showed promising effects of the enhanced Lumen intervention on the primary neural target related to cognitive control—the right dlPFC— and depression and anxiety symptoms.^81^ These results are consistent with the ENGAGE-2 RCT which showed significant effects of an integrated collaborative care intervention combining PST with the Group Lifestyle Balance video program on the right dlPFC in the cognitive control circuit and depressive and anxiety symptoms.^38^ The ENGAGE-1 RCT showed threat-related amygdala activation mediating the effect of in-person PST on depression and problem-solving outcomes.^88^

In addition, the meta-analytic platform Neurosynth^89^ with the search term “threat” was used to define the negative affect network. Analysis of Functional Neuroimaging's (AFNI's) 3dExtrema function was then used to identify peaks corresponding to our a priori regions of interest. Because some terms yielded maps with excessively large spatial extent, we imposed a restriction that each peak have a minimum z-score of 6 and each region extend no farther than 10 mm from the peak. For the amygdala, Neurosynth maps were intersected with anatomically defined boundaries from the Automated Anatomical Labeling atlas.^90^

**Table S1.** **Baseline Characteristics Among Participants with and Without Quality fMRI data at 18 Weeks**

| **Characteristic** | **Participants with fMRI data** | **Participants without fMRI data** | **P value** |
| --- | --- | --- | --- |
|  | **(n=152)** | **(n=48)** |  |
| Group assignment |  |  | 1.00 |
| Lumen | 76 (50) | 24 (50) |  |
| Human | 38 (25) | 12 (25) |  |
| Waitlist | 38 (25) | 12 (25) |  |
| Age, years † | 35.9 ± 11.7 | 38.8 ± 12.4 | 0.15 |
| Female, n (%) † | 115 (75.7) | 40 (83.3) | 0.27 |
| Race/Ethnicity, n (%) † |  |  | 0.69 |
| Non-Hispanic White | 34 (22.4) | 9 (18.8) |  |
| Non-Hispanic Black | 36 (23.7) | 14 (29.2) |  |
| Asian/Pacific Islander | 31 (20.4) | 11 (22.9) |  |
| Hispanic | 46 (30.3) | 11 (22.9) |  |
| Other (e.g., declined to state, multirace) | 5 (3.3) | 3 (6.3) |  |
| Education, n (%) † |  |  | 0.44 |
| High school/GED or less | 12 (7.9) | 7 (14.6) |  |
| College - 1 year to 3 years | 31 (20.4) | 11 (22.9) |  |
| College - 4 years or more | 51 (33.6) | 12 (25) |  |
| Post college | 58 (38.2) | 18 (37.5) |  |
| Income, n (%) |  |  | 0.56 |
| < $35,000 | 43 (28.3) | 15 (31.3) |  |
| $35,000- <$55,000 | 27 (17.8) | 10 (20.8) |  |
| $55,000- <$75,000 | 15 (9.9) | 7 (14.6) |  |
| >=$75,000 | 67 (44.1) | 16 (33.3) |  |
| PHQ-9 category, n (%) † |  |  | 0.58 |
| minimal depression 0-4 | 2 (1.3) | 2 (4.2) |  |
| mild depression 5-9 | 22 (14.5) | 6 (12.5) |  |
| Moderate depression 10-14 | 90 (59.2) | 26 (54.2) |  |
| Moderately severe depression 15-19 | 38 (25) | 14 (29.2) |  |
| GAD-7 category, n (%) † |  |  | 0.11 |
| minimal anxiety 0-4 | 2 (1.3) | 1 (2.1) |  |
| mild anxiety 5-9 | 37 (24.3) | 5 (10.4) |  |
| moderate anxiety 10-14 | 113 (74.3) | 42 (87.5) |  |
| Digital Health Literacy, n (%) † |  |  | 0.37 |
| Low 1-1.999 | 0 (0.0) | 0 (0.0) |  |
| Medium 2-2.999 | 20 (13.2) | 4 (8.3) |  |
| High 3-4 | 132 (86.8) | 44 (91.7) |  |

Abbreviations: GAD-7, Generalized Anxiety Disorder-7; GED, general educational development; PHQ-9, Patient Health Questionnaire-9.

^*^Values are mean ± SD unless noted otherwise.

### †Prognostic factors for randomization: age, sex, race/ethnicity, education, digital health literacy, PHQ-9, and GAD-7.

### Section G: Treatment Effects on Secondary Neural and Patient-Reported Target Measures and Patient-Reported Functional Outcomes

**Table S2. Treatment effects on secondary neural and patient-reported target measures**

|  |  | | |  | **Superiority test** | | | | | | **Noninferiority test** | | |
| --- | --- | --- | --- | --- | --- | --- | --- | --- | --- | --- | --- | --- | --- |
|  | **Unadjusted mean ± SD** | | |  | **Lumen versus Waitlist** | | | **Human versus Waitlist** | | | **Lumen versus Human** | | |
| **Neural Measure** | **Lumen** | **Human** | **Waitlist** | | **Mean difference (95% CI)*** | **P/P_adj_*** | **Cohen's d**†  **(Lumen-Waitlist)** | **Mean difference (95% CI)*** | **P/P_adj_*** | **Cohen's d**† **(Human-Waitlist)** |  |  |  |
| dlPFC L (Cognitive Control Circuit) | n=71 | n=36 | n=34 | |  |  |  |  |  |  |  |  |  |
| baseline | 0.14 ± 0.43 | 0.07 ± 0.41 | 0.02 ± 0.38 | |  |  |  |  |  |  |  |  |  |
| change at 18 weeks | -0.07 ± 0.58 | 0.06 ± 0.47 | 0.00 ± 0.46 | | -0.08 (-0.3, 0.15) | 0.5/0.59 | -0.14 | 0.06 (-0.17, 0.28) | 0.62/0.68 | 0.12 | Not Applicable | | |
| Amygdala R (Non-conscious Negative Affect Circuit) | n=64 | n=30 | n=32 | |  |  |  |  |  |  |  |  |  |
| baseline | 0.02 ± 0.16 | 0.02 ± 0.17 | 0.04 ± 0.19 | |  |  |  |  |  |  |  |  |  |
| change at 18 weeks | 0.01 ± 0.24 | 0.04 ± 0.27 | -0.07 ± 0.27 | | 0.08 (-0.03, 0.19) | 0.15/0.22 | 0.32 | 0.11 (-0.03, 0.24) | 0.13/0.20 | 0.39 | Not Applicable | | |
| Amygdala L (Non-conscious Negative Affect Circuit) | n=64 | n=30 | n=32 | |  |  |  |  |  |  |  |  |  |
| baseline | 0.02 ± 0.17 | 0.01 ± 0.20 | 0.03 ± 0.18 | |  |  |  |  |  |  |  |  |  |
| change at 18 weeks | 0.01 ± 0.29 | 0.05 ± 0.32 | -0.02 ± 0.28 | | 0.03 (-0.09, 0.16) | 0.6/0.68 | 0.11 | 0.07 (-0.08, 0.22) | 0.36/0.47 | 0.23 | Not Applicable | | |
|  | **Unadjusted mean ± SD** | | | | **Lumen versus Waitlist** | | | **Human versus Waitlist** | | | **Lumen versus Human** | | |
| **Patient-reported Target Measure** | **Lumen** | **Human** | **Waitlist** | | **Model-based mean difference (95%CI**)‡ | **P/P_adj_**  **(2-sided)**‡ | **Cohen’s d**† **(Lumen-Waitlist)** | **Model-based mean difference (95%CI)**‡ | **P/P_adj_ (2-sided)**‡ | **Cohen’s d**† **(Human-Waitlist)** | **Model-based mean difference (90%CI)**‡ | **P (1-side)**‡ | **Cohen’s d**† **(Human-Lumen)** |
| Dysfunctional Attitudes Scale (DAS)§ | n=96 | n=48 | n=49 | |  |  |  |  |  |  |  |  |  |
| baseline | 133.8 ± 35.76 | 143.4 ± 33.17 | 140.9 ± 34.24 | |  |  |  |  |  |  |  |  |  |
| change at 18 weeks | -14.7 ± 29.02 | -20.5 ± 40.92 | -11.1 ± 24.40 | | -5.48 (-15.45, 4.49) | 0.28/0.37 | 0.13 | -7.95 (-19.47, 3.57) | 0.17/0.24 | 0.28 | 2.47 (-5.97, 10.92) | 0.36 | 0.17 |
| Penn State Worry Questionnaire (PSWQ)¶ | n=97 | n=48 | n=49 | |  |  |  |  |  |  |  |  |  |
| baseline | 59.99 ± 11.21 | 59.96 ± 11.19 | 61.44 ± 11.35 | |  |  |  |  |  |  |  |  |  |
| change at 18 weeks | -8.35 ± 12.87 | -9.38 ± 15.52 | -2.61 ± 10.62 | | -6.23 (-10.32, -2.14) | 0.003/0.01 | 0.47 | -6.99 (-11.72, -2.25) | 0.004/0.02 | 0.51 | 0.75 (-2.7, 4.2) | 0.39 | 0.07 |
| Positive and Negative Affect Schedule (PANAS)‖ |  |  |  | |  |  |  |  |  |  |  |  |  |
| Positive | n=97 | n=48 | n=49 | |  |  |  |  |  |  |  |  |  |
| baseline | 26.90 ± 6.96 | 26.10 ± 6.07 | 26.26 ± 7.44 | |  |  |  |  |  |  |  |  |  |
| change at 18 weeks | 6.04 ± 7.55 | 6.46 ± 8.01 | 3.61 ± 7.03 | | 2.47 (0.03, 4.91) | 0.048/0.09 | 0.33 | 2.69 (-0.14, 5.52) | 0.06/0.11 | 0.38 | -0.23 (-2.29, 1.83) | 0.44 | 0.05 |
| Negative | n=97 | n=48 | n=49 | |  |  |  |  |  |  |  |  |  |
| baseline | 27.22 ± 7.39 | 27.86 ± 7.17 | 26.68 ± 7.24 | |  |  |  |  |  |  |  |  |  |
| change at 18 weeks | -4.04 ± 7.66 | -7.19 ± 9.05 | -0.22 ± 7.47 | | -3.47 (-5.97, -0.97) | 0.007/0.03 | 0.50 | -6.24 (-9.15, -3.34) | <.001/<.001 | 0.84 | 2.78 (0.67, 4.89) | 0.14 | 0.39 |

Abbreviations: CI, confidence interval; DAS, Dysfunctional Attitudes Scale; PANAS, Positive and Negative Affect Schedule; PSWQ, Penn State Worry Questionnaire.

**t* tests. Both P values and FDR-adjusted P values were reported.

†The mean difference between two groups divided by the pooled standard deviation.

‡Model-based mean differences with 95% CI and P and FDR-adjusted P values (2-sided) for superiority tests and with 90% CI and P values (1-sided) for noninferiority tests were generated from ordinary least square regression models, adjusted for baseline value of the outcome of interest.

§DAS (Form A) is a 40-item self-reported scale that measures the presence and intensity of dysfunctional attitudes. Each item is rated a 7-point Likert scale (7 = fully agree; 1 = fully disagree). The higher the sum of the 40-items, the more dysfunctional attitudes an individual possesses.

¶PSWQ is a self-reported, 16-item, Likert-type scale that measures the trait of worry. Each item is related on a 1-5 scale. The total score of the scale ranges from 16 to 80, with higher scores indicating higher trait worry.

‖PANAS consists of two 10-item self-reported scales to measure positive and negative affect. Each item asks about the extent one has felt a positive or negative feeling on a 5-point scale of 1 (not at all) to 5 (very much). Positive affect scores can range from 10 – 50, with higher scores representing higher levels of positive affect. Negative affect scores can range from 10 – 50, with lower scores representing lower levels of negative affect.

**Table S3. Per-Protocol non-inferiority tests for Lumen versus Human among participants completing all 8 PST sessions**

|  | **Unadjusted mean ± SD** | | **Per-Protocol** | | |
| --- | --- | --- | --- | --- | --- |
| **Symptom** | **Luman** | **Human** | **Model-based mean difference (90%CI)*** | **P (1-sided)*** | **Cohen’s d**† **(Human-Lumen)** |
| **Patient-reported target measures** |  |  |  |  |  |
| SPSI-R:S raw score‡ | n=73 | n=44 |  |  |  |
| Baseline | 12.22 ± 2.87 | 11.29 ± 2.71 | 0.04 (-0.73, 0.81) | 0.53 | 0.12 |
| Change at 18 weeks | 1.53 ± 2.61 | 1.84 ± 2.61 |  |  |  |
| PPO raw score‡ | n=73 | n=44 |  |  |  |
| Baseline | 11.25 ± 3.85 | 10.34 ± 3.71 | -0.08 (-1.17, 1.02) | 0.46 | 0.11 |
| Change at 18 weeks | 2.08 ± 3.4 | 2.5 ± 4.17 |  |  |  |
| NPO raw score‡ | n=73 | n=44 |  |  |  |
| Baseline | 9.32 ± 4.68 | 10.09 ± 3.98 | 0.22 (-1, 1.43) | 0.41 | 0.13 |
| Change at 18 weeks | -2.52 ± 4.21 | -3.07 ± 4.29 |  |  |  |
| RPS raw score‡ | n=73 | n=44 |  |  |  |
| Baseline | 11.34 ± 4.53 | 10.02 ± 4.31 | 0.65 (-0.57, 1.88) | 0.73 | -0.0006 |
| Change at 18 weeks | 1.62 ± 4.38 | 1.61 ± 4.45 |  |  |  |
| ICS raw score‡ | n=73 | n=44 |  |  |  |
| Baseline | 5.26 ± 4 | 6.23 ± 3.96 | -0.46 (-1.55, 0.63) | 0.69 | 0.02 |
| Change at 18 weeks | -0.38 ± 4.07 | -0.48 ± 4.16 |  |  |  |
| AS raw score‡ | n=73 | n=44 |  |  |  |
| Baseline | 6.9 ± 4.41 | 7.61 ± 4.91 | 0.17 (-0.97, 1.31) | 0.42 | 0.12 |
| Change at 18 weeks | -1.07 ± 3.97 | -1.55 ± 4.27 |  |  |  |
| Dysfunctional Attitudes Scale (DAS)§ | n=73 | n=44 |  |  |  |
| Baseline | 133.52 ± 35.11 | 143.64 ± 34.21 | -0.25 (-9.76, 9.27) | 0.51 | 0.14 |
| Change at 18 weeks | -16.26 ± 29.29 | -21.27 ± 42.28 |  |  |  |
| Penn State Worry Questionnaire (PSWQ)¶ | n=73 | n=44 |  |  |  |
| Baseline | 60.64 ± 11.29 | 60.93 ± 11.22 | -0.78 (-4.69, 3.12) | 0.60 | -0.04 |
| Change at 18 weeks | -10.11 ± 12.71 | -9.5 ± 15.97 |  |  |  |
| Positive and Negative Affect Schedule (PANAS)‖ |  |  |  |  |  |
| Positive | n=73 | n=44 |  |  |  |
| Baseline | 26.1 ± 6.82 | 26.07 ± 6.39 | 0.15 (-2.21, 2.5) | 0.53 | -0.02 |
| Change at 18 weeks | 6.86 ± 7.74 | 6.73 ± 8.11 |  |  |  |
| Negative | n=73 | n=44 |  |  |  |
| Baseline | 27.1 ± 7.56 | 27.93 ± 7.5 | 2.42 (0.17, 4.68) | 0.16 | 0.34 |
| Change at 18 weeks | -4.7 ± 8.26 | -7.66 ± 9.13 |  |  |  |
| **Patient-reported outcomes** |  |  |  |  |  |
| HADS_Total** | n=73 | n=44 |  |  |  |
| Baseline | 18.44 ± 5.03 | 18.91 ± 4.72 | 0.63 (-1.47, 2.73) | 0.35 | 0.13 |
| Change at 18 weeks | -6.45 ± 6.85 | -7.41 ± 8.35 |  |  |  |
| HADS_Depression†† | n=73 | n=44 |  |  |  |
| Baseline | 7.16 ± 3.01 | 7.57 ± 3.11 | 0.25 (-0.83, 1.33) | 0.38 | 0.13 |
| Change at 18 weeks | -2.67 ± 3.4 | -3.18 ± 4.66 |  |  |  |
| HADS_Anxiety†† | n=73 | n=44 |  |  |  |
| Baseline | 11.27 ± 3.42 | 11.34 ± 3.15 | 0.4 (-0.84, 1.65) | 0.34 | 0.10 |
| Change at 18 weeks | -3.78 ± 4.39 | -4.23 ± 4.57 |  |  |  |
| Sheehan Disability Scale (SDS)‡‡ | n=73 | n=44 |  |  |  |
| Baseline | 18.48 ± 6.39 | 18.39 ± 6.88 | 0.82 (-1.69, 3.34) | 0.34 | 0.08 |
| Change at 18 weeks | -6.47 ± 8.59 | -7.23 ± 9.87 |  |  |  |
| Work productivity and activity impairment questionnaire (WPAI)§§ |  |  |  |  |  |
| Percent work time missed | n=60 | n=35 |  |  |  |
| Baseline | 0.06 ± 0.11 | 0.13 ± 0.2 | 0.03 (-0.03, 0.1) | 0.26 | 0.47 |
| Change at 18 weeks | 0.02 ± 0.19 | -0.07 ± 0.22 |  |  |  |
| Percent impairment at work | n=60 | n=35 |  |  |  |
| Baseline | 0.28 ± 0.25 | 0.28 ± 0.28 | 0.03 (-0.05, 0.11) | 0.31 | 0.13 |
| Change at 18 weeks | -0.1 ± 0.26 | -0.14 ± 0.34 |  |  |  |
| Percent overall work productivity loss | n=60 | n=35 |  |  |  |
| Baseline | 0.31 ± 0.27 | 0.34 ± 0.33 | 0.04 (-0.05, 0.14) | 0.29 | 0.24 |
| Change at 18 weeks | -0.07 ± 0.31 | -0.16 ± 0.39 |  |  |  |
| Percent activity impairment | n=73 | n=43 |  |  |  |
| Baseline | 0.33 ± 0.27 | 0.36 ± 0.28 | 0.06 (-0.01, 0.13) | 0.21 | 0.25 |
| Change at 18 weeks | -0.11 ± 0.28 | -0.19 ± 0.33 |  |  |  |

Abbreviations: AS, avoidant problem-solving style; DAS, Dysfunctional Attitudes Scale; HADS, Hospital Anxiety and Depression Scale; ICS, impulsive/careless problem-solving style; NPO, negative problem orientation; PANAS, Positive and Negative Affect Schedule; PPO, positive problem orientation; PSWQ, Penn State Worry Questionnaire; RPS, rational problem-solving style; SDS, Sheehan Disability Scale; SPSI-R:S, Social Problem-solving Index-Revised Short Form; WPAI, Work productivity and activity impairment questionnaire.

*Model based mean difference (95%/90% CI) and p value/one-sided p value were generated from regression model adjusted for baseline value of the interest.

†The mean difference between two groups divided by the pooled standard deviation.

‡SPSI-R:S score=(PPO raw score/5)+(20- NPO raw score)/5+ (RPS raw score/5)+(20- ICS raw score)/5+(20- AS raw score)/5; the higher the score the more productive overall problem-solving orientation and skills. Subscales (PPO, NPO, RPS, ICS, and AS) are raw scores without reversal.

§DAS (Form A) is a 40-item self-reported scale that measures the presence and intensity of dysfunctional attitudes. Each item is rated a 7-point Likert scale (7 = fully agree; 1 = fully disagree). The higher the sum of the 40-items, the more dysfunctional attitudes an individual possesses.

¶PSWQ is a self-reported, 16-item, Likert-type scale that measures the trait of worry. Each item is related on a 1-5 scale. The total score of the scale ranges from 16 to 80, with higher scores indicating higher trait worry.

‖PANAS consists of two 10-item self-reported scales to measure positive and negative affect. Each item asks about the extent one has felt a positive or negative feeling on a 5-point scale of 1 (not at all) to 5 (very much). Positive affect scores can range from 10 – 50, with higher scores representing higher levels of positive affect. Negative affect scores can range from 10 – 50, with lower scores representing lower levels of negative affect.

**HADS, including 7 questions for anxiety and 7 questions for depression, measures self-reported anxiety and depression in a general medical population of patients. Each item on the questionnaire is scored from 0-3. Total score for the entire scale (emotional distress) ranges from 0 to 42, with higher score indicating more distress.

††Scores range from 0-21, with 0-7 = Normal; 8-10 = Borderline abnormal (borderline case); 11-21 = Abnormal (case).

‡‡SDS is a validated questionnaire that measures functional disability. Patients rate the extent to which their symptoms impair work/school, social, and family life on a visual analog scale from 0 to 10 and answer the number of days when their symptoms cause them to miss work/school and be unproductive at work/school. Total scores range 0-30, with higher scores indicating greater functional impairment.

§§WPAI was created as a patient-reported quantitative assessment of the amount of absenteeism, presenteeism and daily activity impairment attributable to general health in the past 7 days.

**Table S4. Treatment effects on patient-reported functional outcomes**

|  |  | | |  | **Superiority test** | | | | | | **Noninferiority test** | | |
| --- | --- | --- | --- | --- | --- | --- | --- | --- | --- | --- | --- | --- | --- |
|  | **Unadjusted mean ± SD** | | |  | **Lumen versus Waitlist** | | | **Human versus Waitlist** | | | **Lumen versus Human** | | |
| **Measure** | **Lumen** | **Human** | **Waitlist** | | **Model-based mean difference (95%CI**)* | **P/P_adj_**  **(2-sided)*** | **Cohen’s d**† **(Lumen-Waitlist)** | **Model-based mean difference (95%CI)*** | **P/P_adj_ (2-sided)*** | **Cohen’s d**† **(Human-Waitlist)** | **Model-based mean difference (90%CI)*** | **P (1-side*** | **Cohen’s d**† **(Human-Lumen)** |
| Sheehan Disability Scale (SDS)‡ | n=96 | n=48 | n=49 | |  |  |  |  |  |  |  |  |  |
| baseline | 17.81 ± 6.75 | 18.62 ± 6.73 | 19.42 ± 6.74 | |  |  |  |  |  |  |  |  |  |
| change at 18 weeks | -5.02 ± 9.17 | -7.40 ± 9.52 | -0.86 ± 7.84 | | -5.08 (-7.77, -2.39) | <.001/<.001 | 0.48 | -7.1 (-10.2, -4) | <.001/<.001 | 0.75 | 2.02 (-0.24, 4.28) | 0.19 | 0.26 |
| Work productivity and activity impairment questionnaire (WPAI)§ |  |  |  | |  |  |  |  |  |  |  |  |  |
| Percent work time missed | n=79 | n=39 | n=40 | |  |  |  |  |  |  |  |  |  |
| baseline | 6.86 ± 12.22 | 11.66 ± 19.41 | 6.46 ± 14.06 | |  |  |  |  |  |  |  |  |  |
| change at 18 weeks | 0.57 ± 18.37 | -6.19 ± 20.55 | 0.02 ± 19.48 | | 0.56 (-5.16, 6.28) | 0.85/0.89 | 0.03 | -1.22 (-7.92, 5.48) | 0.72/0.77 | 0.31 | 0.02 (-0.03, 0.07) | 0.33 | 0.35 |
| Percent impairment at work | n=79 | n=39 | n=40 | |  |  |  |  |  |  |  |  |  |
| baseline | 25.47 ± 24.29 | 28.84 ± 27.19 | 33.41 ± 25.24 | |  |  |  |  |  |  |  |  |  |
| change at 18 weeks | -9.11 ± 27.28 | -12.31 ± 32.88 | -0.75 ± 27.77 | | -11.87 (-20.57, -3.17) | 0.008/0.03 | 0.30 | -12.81 (-22.87, -2.75) | 0.01/0.03 | 0.38 | 0.01 (-0.06, 0.08) | 0.43 | 0.11 |
| Percent overall work productivity loss | n=79 | n=39 | n=40 | |  |  |  |  |  |  |  |  |  |
| baseline | 29.23 ± 26.77 | 34.15 ± 31.4 | 35.99 ± 27.16 | |  |  |  |  |  |  |  |  |  |
| change at 18 weeks | -7.9 ± 31.27 | -13.87 ± 38.09 | 0.55 ± 34.36 | | -11.7 (-21.59, -1.81) | 0.02/0.04 | 0.26 | -13.21 (-24.66, -1.76) | 0.02/0.04 | 0.40 | 0.02 (-0.07, 0.1) | 0.41 | 0.18 |
| Percent activity impairment | n=96 | n=47 | n=49 | |  |  |  |  |  |  |  |  |  |
| baseline | 32.3 ± 26.13 | 36.6 ± 28.04 | 38.8 ± 26.93 | |  |  |  |  |  |  |  |  |  |
| change at 18 weeks | -11.04 ± 30 | -17.66 ± 32.85 | -4.69 ± 26.54 | | -9.87 (-18.13, -1.62) | 0.02/0.04 | 0.22 | -14.77 (-24.34, -5.19) | 0.003/0.01 | 0.44 | 0.05 (-0.02, 0.12) | 0.23 | 0.21 |

Abbreviations: CI, confidence interval; SDS, Sheehan Disability Scale; WPAI, Work productivity and activity impairment questionnaire.

*Model-based mean differences with 95% CI and P and FDR-adjusted P values (2-sided) for superiority tests and with 90% CI and P values (1-sided) for noninferiority tests were generated from ordinary least square regression models, adjusted for baseline value of the outcome of interest.

†The mean difference between two groups divided by the pooled standard deviation.

‡SDS is a validated questionnaire that measures functional disability. Patients rate the extent to which their symptoms impair work/school, social, and family life on a visual analog scale from 0 to 10 and answer the number of days when their symptoms cause them to miss work/school and be unproductive at work/school. Total scores range 0-30, with higher scores indicating greater functional impairment.

§WPAI was created as a patient-reported quantitative assessment of the amount of absenteeism, presenteeism and daily activity impairment attributable to general health in the past 7 days.

### Section H: Associations between Problem-Solving Ability and Clinical Symptom Outcomes (Path B)

### Table S5. Associations between changes in overall problem-solving ability and in clinical symptoms (Path B)*†‡

| **Clinical symptoms** |  | **Lumen vs. Waitlist** | | **Human vs. Waitlist** | |
| --- | --- | --- | --- | --- | --- |
|  |  | **Estimate (95% CI)** | **P value** | **Estimate (95% CI)** | **P value** |
| **Psychological distress** | **Main effect** | -0.76 (-1.46, -0.07) | 0.03 | -0.84 (-1.56, -0.13) | 0.02 |
|  | **Interaction** | -0.12 (-0.94, 0.69) | 0.77 | -0.57 (-1.56, 0.42) | 0.26 |
| **Depression** | **Main effect** | -0.26 (-0.61, 0.10) | 0.15 | -0.28 (-0.69, 0.12) | 0.17 |
|  | **Interaction** | -0.17 (-0.58, 0.25) | 0.43 | -0.51 (-1.07, 0.04) | 0.07 |
| **Anxiety** | **Main effect** | -0.53 (-0.98, -0.08) | 0.02 | -0.57 (-0.99, -0.15) | 0.01 |
|  | **Interaction** | 0.08 (-0.45, 0.60) | 0.77 | -0.03 (-0.61, 0.55) | 0.93 |

Abbreviations: AS, avoidant problem-solving style; CI, confidence interval; HADS, Hospital Anxiety and Depression Scale;

ICS, impulsive/careless problem-solving style; NPO, negative problem orientation; PPO, positive problem orientation; RPS, rational problem-solving style; SPSI-R:S, Social Problem-solving Index-Revised Short Form.

*****SPSI-R:S total score=(PPO score/5)+(20- NPO score)/5+ (RPS score/5)+(20- ICS score)/5+(20- AS score)/5; the higher the score the stronger the overall problem-solving ability.

### †HADS, including 7 questions for anxiety and 7 questions for depression, measures self-reported anxiety and depression in a general medical population of patients. Each item on the questionnaire is scored from 0-3. Total score for the entire scale (emotional distress) ranges from 0 to 42, with higher score indicating more distress.

‡For each HADS outcome, the association between change in overall problem-solving ability and change in symptom severity was tested in an ANCOVA model, with the HADS change score as the dependent variable, change in problem-solving ability and its interaction with treatment assignment as predictors, and baseline HADS score as a covariate.

### Section I: Adverse Events Summary

**Table S6. Summary of adverse events by body system**

| **System** |  | **Total** | |
| --- | --- | --- | --- |
|  |  | ***Serious*** | ***Not Serious*** |
| Cardiovascular | **Unexpected and related** | 0 | 0 |
|  | **Unexpected and unrelated** | 0 | 0 |
|  | **Expected and related** | 0 | 0 |
|  | **Expected and unrelated** | 0 | 1 |
| Pulmonary/  Respiratory | **Unexpected and related** | 0 | 0 |
|  | **Unexpected and unrelated** | 0 | 0 |
|  | **Expected and related** | 0 | 0 |
|  | **Expected and unrelated** | 1 | 4 |
| Hematological | **Unexpected and related** | 0 | 0 |
|  | **Unexpected and unrelated** | 0 | 0 |
|  | **Expected and related** | 0 | 0 |
|  | **Expected and unrelated** | 0 | 0 |
| Metabolic | **Unexpected and related** | 0 | 0 |
|  | **Unexpected and unrelated** | 0 | 0 |
|  | **Expected and related** | 0 | 0 |
|  | **Expected and unrelated** | 0 | 0 |
| Musculoskeletal | **Unexpected and related** | 0 | 0 |
|  | **Unexpected and unrelated** | 0 | 4 |
|  | **Expected and related** | 0 | 0 |
|  | **Expected and unrelated** | 0 | 9 |
| Hepatobiliary | **Unexpected and related** | 0 | 0 |
|  | **Unexpected and unrelated** | 0 | 0 |
|  | **Expected and related** | 0 | 0 |
|  | **Expected and unrelated** | 0 | 0 |
| Reproductive | **Unexpected and related** | 0 | 0 |
|  | **Unexpected and unrelated** | 0 | 1 |
|  | **Expected and related** | 0 | 0 |
|  | **Expected and unrelated** | 0 | 4 |
| Neurological | **Unexpected and related** | 0 | 0 |
|  | **Unexpected and unrelated** | 0 | 0 |
|  | **Expected and related** | 0 | 0 |
|  | **Expected and unrelated** | 0 | 1 |
| Gastrointestinal | **Unexpected and related** | 0 | 0 |
|  | **Unexpected and unrelated** | 0 | 0 |
|  | **Expected and related** | 0 | 0 |
|  | **Expected and unrelated** | 0 | 1 |
| Psychological | **Unexpected and related** | 0 | 0 |
|  | **Unexpected and unrelated** | 0 | 0 |
|  | **Expected and related** | 0 | 0 |
|  | **Expected and unrelated** | 0 | 2 |
| Renal/  Urologic | **Unexpected and related** | 0 | 0 |
|  | **Unexpected and unrelated** | 0 | 0 |
|  | **Expected and related** | 0 | 0 |
|  | **Expected and unrelated** | 0 | 2 |
| More than one | **Unexpected and related** | 0 | 0 |
|  | **Unexpected and unrelated** | 0 | 0 |
|  | **Expected and related** | 0 | 0 |
|  | **Expected and unrelated** | 0 | 2 |
| Other | **Unexpected and related** | 0 | 0 |
|  | **Unexpected and unrelated** | 0 | 0 |
|  | **Expected and related** | 0 | 0 |
|  | **Expected and unrelated** | 0 | 5 |
| **Overall** | **Unexpected and related** | 0 | 0 |
|  | **Unexpected and unrelated** | 0 | 5 |
|  | **Expected and related** | 0 | 0 |
|  | **Expected and unrelated** | 1 | 30 |
|  | **GRAND TOTAL (N=36*)** | 1 | 35 |

***** This table only includes events reported as part of regular surveillance among randomized participants.
